# When Does IPCW Help? Simulation and Real-World Evidence on Censoring Adjustment in Survival Analysis

**DOI:** 10.1101/2025.08.22.25334253

**Authors:** Hsin Yi Chen, Tara V. Anand, Linying Zhang, George Hripcsak

## Abstract

Estimating treatment effects from time-to-event data in observational studies requires careful adjustment for both confounding and informative censoring. While inverse probability of treatment weighting (IPTW) and inverse probability of censoring weighting (IPCW) have been used to address these sources of bias separately, their combined application remains underexplored, especially in high-dimensional, real-world datasets. In this paper, we benchmark IPTW, IPCW, and their combination to estimate survival curves, restricted mean survival time (RMST), and hazard ratios (HR). Our simulation studies vary strengths of informative censoring and introduce non-proportional hazards, while our real-world study uses a large-scale electronic health record (EHR) dataset (~ 50,000 covariates and >40,000 patients). Our simulations showed that IPCW reduces survival curve estimation error in the presence of informative censoring, but only reduces HR and RMST bias when the strength of informative censoring additionally differs by treatment group. In our real-world study, IPTW alone was typically sufficient for HR estimation, suggesting that when confounding is the primary source of bias and well-addressed through large-scale adjustment, censoring adjustment may yield limited additional benefit. Ultimately, the utility of IPCW likely depends on the underlying data-generating process, the relative magnitude of censoring bias, and the estimand of interest.

## Introduction

Estimating treatment effects of drugs on health outcomes is a central goal in both randomized controlled trials (RCTs) and observational studies. In settings where the outcome of interest is a time-to-event (i.e., survival outcome), the data is also frequently subject to right-censoring, where the event of interest (e.g., death, disease onset) is not observed within the study period. Censoring arises due to various reasons: in RCTs, censoring may be due to administrative end-of-study truncation or participant dropout, while in the observational data setting (e.g., using real-world data sources such as electronic health records (EHRs) or administrative claims), censoring may result from patients changing healthcare providers, missing follow-up visits, or losing insurance coverage.

Statistical survival analysis methods, such as the Kaplan-Meier (KM) curve estimator ^16^ and the Cox Proportional Hazards (PH) model ^7^, rely on the assumption of non-informative censoring; i.e., censoring is independent of the outcome of interest conditional on observed covariates. In practice, however, this assumption is difficult to verify, and when violated, it can lead to biased estimates because the censored population may systematically differ from the uncensored population in ways related to the outcome risk ^15,18,19,23^.

To address informative censoring, inverse probability of censoring weighting (IPCW) has been proposed ^24,26^. IPCW corrects for informative censoring by upweighting those who remain in the risk set but have similar covariate profiles to those who are censored. This creates a pseudopopulation that would have been observed had censoring not occurred. Conceptually, this is similar to inverse probability of treatment weighting (IPTW) ^27^, which adjusts for confounding by weighting patients based on the probability of their treatment assignment, thus creating a pseudopopulation in which measured confounders are equally distributed across the treatment groups. While the theoretical foundations for IPCW and IPTW are well established, their joint application to adjust for both confounding and censoring bias in the context of high-dimensional, real-world survival data remains underexplored.

In this paper, we examine IPTW, IPCW, and their combination, for estimating treatment effects in real-world health data with high-dimensional covariates and right-censored survival outcomes. Our contributions are as follows:

1. We conduct a simulation study to benchmark the performance of IPTW, IPCW, and their combination across a range of data-generating scenarios. We vary the strength of informative censoring and consider violations of the proportional hazards assumption in the presence of informative censoring. Then, we evaluate the performance of the weights on multiple treatment effect estimands, including hazard ratios (HR), differences in restricted mean survival time (RMST), and Kaplan-Meier survival curves.
2. We apply these weighting methods to real-world observational data from a large EHR database containing almost 50,000 covariates and over 40,000 patients, to estimate the comparative effect of ACE inhibitors (ACEi) versus thiazide diuretics on risk of acute myocardial infarction (AMI).
3. We provide practical recommendations for when to use IPCW, IPTW, and their combination in high-dimensional survival settings.

### Related work

A substantial body of literature has explored methods for estimating average treatment effects (ATE) using both statistical and machine learning approaches. Foundational work in statistics laid the groundwork for causal inference with observational data, introducing frameworks such as the potential outcomes model and marginal structural models ^24,28,29^. More recently, machine learning-based methods such as Targeted Maximum Likelihood Estimation (TMLE) ^34,35^, Bayesian Additive Regression Trees (BART) ^6,14^, causal forests ^12,37^, and double machine learning ^5^ have been developed to improve ATE estimation in flexible, high-dimensional settings. However, these studies primarily focus on ATEs in the non-survival setting, while researchers in healthcare are often interested in right-censored, time-to-event outcomes.

Recent work has adapted modern machine-learning estimators to survival data by modeling both the event process and the censoring mechanism. Rytgaard and van der Laan ^30^ extended TMLE to accommodate survival outcomes, offering robust estimators of treatment-specific survival probabilities and restricted mean survival time (RMST) even under informative censoring or non-proportional hazards. This work focused on treatment-specific survival probabilities as a causal parameter and avoided the Cox proportional hazards ratio (HR) entirely, which aligns with the growing body of literature that cautions against the use of the hazards ratio as a causal parameter ^9,13^. Despite its limitations, HR remains one of the most common estimands in clinical and epidemiological literature due to its interpretability and widespread familiarity. Our work acknowledges these ongoing discussions and includes HR alongside RMST and survival curves to provide a comprehensive comparison of treatment effect estimands under various adjustment strategies.

Inverse Probability of Censoring Weighting (IPCW) addresses informative censoring by modeling the censoring process and applying the resulting weights to otherwise standard estimators. This method has been applied to a number of methods, including KM curves and log rank tests ^25^, Cox PH models ^3,25^, win ratios ^8,22^, and restricted mean survival times ^41^. Applications of these IPCW methods in clinical settings, however, are typically limited to randomized trials ^39^ or low-dimensional settings with predefined clinical variables ^21,38^. These clinical studies also often focus on a single survival estimand (e.g., HR or Kaplan-Meier curve). Our work extends this line of research by exploring IPCW in a high-dimensional EHR setting with almost 50,000 baseline covariates and evaluating multiple estimands.

Importantly, IPCW adjustment does not adjust for confounding, another common source of bias in observational studies. Thus, combining IPCW with inverse probability of treatment weighting (IPTW) has been proposed to simultaneously adjust for informative censoring and confounding ^3,31^. However, few studies have systematically examined the performance of IPCW, IPTW, and their combination across estimands or under high-dimensional conditions. Our work fills this gap by benchmarking all three approaches using both simulation studies and a large-scale EHR dataset. We provide empirical evidence and practical guidance on their relative performance and use cases.

In summary, our work is among the first to benchmark IPCW, IPTW, and their combination in high-dimensional survival settings, examine their effects across multiple estimands, and validate performance in a large-scale, high-dimensional real-world EHR dataset.

## Background

We begin by reviewing censoring definitions, followed by a brief review of both the Kaplan-Meier survival curve and the Cox proportional hazards model as they are two commonly used models in survival analysis and widely used in clinical research.

### Non-informative Censoring Definition

In the survival analysis literature, there exist several assumptions about the censoring mechanism, including random censoring, independent censoring, and non-informative censoring. Although these definitions are not synonymous, they are often used interchangeably. Thus, we explicitly define *non-informative censoring* here, using definitions proposed by Kleinbaum and Klein ^17^.

Formally, let *T* denote the true event time and *C* denote the censoring time. The observed time for each individual is 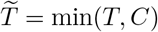 with an indicator variable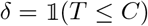, where *δ* = 1 indicates an observed event, and *δ* = 0 indicates censoring.

Recall that the hazard function, or instantaneous event rate of a non-negative random variable *U*, is

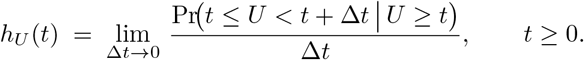

Intuitively, *h*_*U*_ (*t*)d*t* approximates the probability that the event occurs in the next infinitesimal interval [*t, t* + Δ*t*), given survival up to *t*.

#### Non-informative censoring (NIC)

Censoring is non-informative if, at every time *t*, the event hazard is unaffected by the fact that the individual has not yet been censored, i.e.,

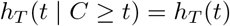

and equivalently, the hazard of censoring *h*_*C*_ is unaffected by having survived past time t, i.e.,

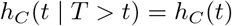

In other words, censored individuals have the same survival distribution as those who remain under observation. An equivalent shorthand for this assumption is written as 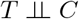.

When 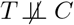 censoring is considered to be informative. However, informative censoring is not always differential. Because we will primarily be studying treatment effect estimation in this paper, we will additionally define *differential* informative censoring:

#### *Differential* informative censoring

Censoring is differentially informative when (1) the hazard of censoring differs by treatment group and (2) within at least one treatment group, the censoring process is informative. Formally, let *Z* ∈{0, 1} denote treatment, *T* denote event time, and *C* denote censoring time.

Differential censoring means that the censoring hazard differs across treatment groups, i.e., there exists some *t* such that

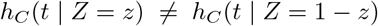

Informative censoring means that, within at least one treatment group *Z*, knowledge of whether a subject will remain uncensored at time *t* changes the hazard of the event, i.e.,

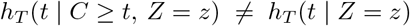

When both conditions hold simultaneously, censoring is *differentially informative* (Figure 1).

**Figure 1.**
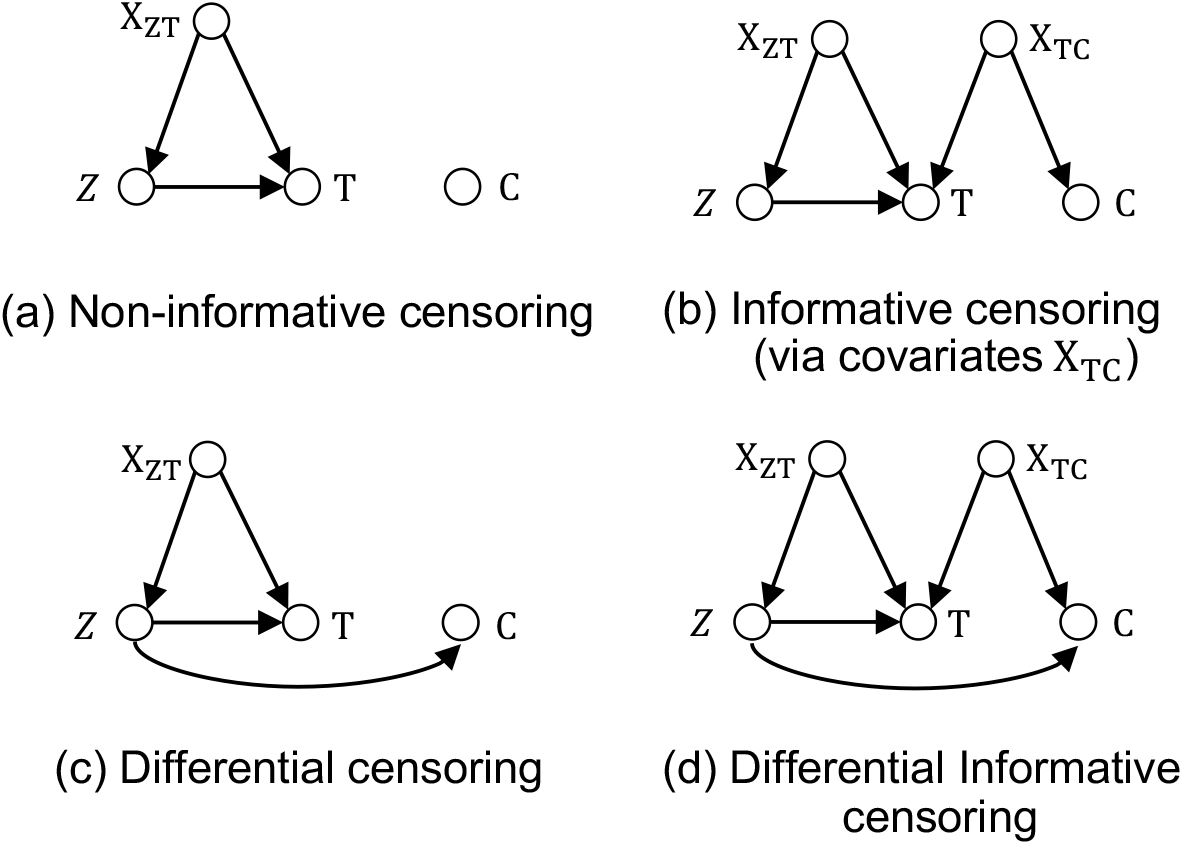
Causal diagram illustrating different types of censoring mechanisms, where *Z* is the treatment, *T* is the outcome time, *C* is the censoring time, and *X* are a set of covariates which can affect treatment, outcome, and/or censoring. (a) Non-informative censoring: the censoring process is independent of the event process given treatment and covariates. (b) Informative (but non-differential) censoring: censoring depends on covariates *X*_*TC*_, which also affects the event, but the censoring mechanism does not differ across treatment groups. (c) Differential (but not informative) censoring: the censoring hazard differs across treatment groups, but there are no covariates that are informative for both the event hazard *and* the censoring hazard. While in this case, censoring is associated with outcome through treatment, this dependence vanishes once we condition on *Z* in treatment effect estimation, so it is not considered informative. (d) Differential informative censoring: both conditions in panels (b) and (c) hold: the hazard of censoring differs by treatment group and, within at least one treatment arm, censoring is informative for the event hazard.

Practically, informative censoring may be relevant in scenarios where certain factors influence health outcomes but are not included in treatment decision-making. This is a situation that may arise when considering factors such as social determinants of health (SDOH). For example, if patient’s follow-up visits or continuity of care are influenced by socioeconomic factors (e.g., housing stability, income, transportation barriers), patients from disadvantaged backgrounds may be more likely to be censored due to disrupted care. If these same SDOH factors also affect health outcomes independently of treatment assignment, then censoring becomes informative. Moreover, if the treatment itself influences patients’ engagement with care (e.g., one treatment has more side effects or requires more frequent monitoring), then this would creates differential informative censoring.

### Kaplan-Meier Estimator

The Kaplan-Meier (KM) estimator is a nonparametric method for estimating survival probabilities over time. The Kaplan-Meier estimator estimates the survival function *S*(*t*), the probability of surviving beyond time *t*. It is written as

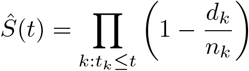

where *t*_*k*_ are the ordered event times, *d*_*k*_ is the number of events (e.g., deaths) at time *t*_*k*_, and *n*_*k*_ is the number of individuals at risk just before time *t*_*k*_.

#### Kaplan-Meier Survival Curve and censoring

The Kaplan-Meier (KM) survival curve accounts for censoring by updating the survival probability only at observed event times. The KM estimator assumes that censoring is non-informative, and the KM estimator will overestimate survival if individuals at higher risk are more likely to be censored. Conversely, if lower-risk individuals are preferentially censored, the KM estimator may underestimate survival ^4^.

### Cox Proportional Hazards Model

The Cox Proportional Hazards (PH) Model is a semi-parametric model consisting of two parts: (1) the underlying baseline hazard function, *λ*_0_(*t*), and (2) the effect parameters, which describe the relationship between the hazards and the covariates:

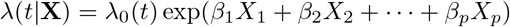

Here,

- *λ*(*t*|**X**) is the hazard function at time *t* given covariates **X** = (*X*_1_, *X*_2_, …, *X*_*p*_)
- *λ*_0_(*t*) is the baseline hazard function, representing the hazard when all covariates are set to zero
- *β*_1_, *β*_2_, …, *β*_*p*_ are the coefficients for the covariates *X*_1_, *X*_2_, …, *X*_*p*_. Each coefficient *β*_*j*_ represents the log hazard ratio for a one-unit increase in *X*_*j*_, holding all other variables constant.

The Cox model allows us to quantify the relative effect of each factor on the time-to-event outcome through the hazard ratio, *exp*(*β*). In order to estimate *β*, this method maximizes the log of the Cox partial likelihood:

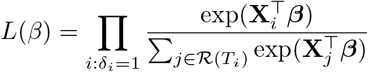

The product is over individuals *i* who experienced the event (i.e., : *δ*_*i*_ = 1). For each individual *i* who experienced the event, the denominator sums over all individuals *j* in the risk set at time *T*_*i*_, the observed time event for individual *i*, i.e. *j* ∈*R*(*T*_*i*_).

The log-partial likelihood can then be written as:

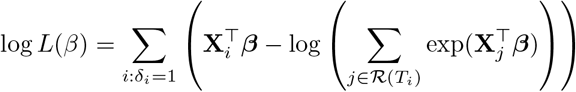

Estimating regression coefficients *β* through maximizing the log partial likelihood relies only on the relative order of event times rather than their exact values. This enables the estimation of regression coefficients, and thus the hazard ratios, without having to make parametric assumptions about the *λ*_0_(*t*).

#### Cox regression and censoring

Censored individuals contribute to the denominator (risk set) of the Cox partial likelihood but do not contribute directly to the numerator since their event times are unknown. If the assumption of non-informative censoring is violated, the resulting hazard ratio estimates may be biased.

## Methods

Next, we present our problem setup, notations, and assumptions, before introducing inverse probability of censoring weighting (IPCW), a method that also involves modeling survival time to address censoring bias. We then discuss inverse probability of treatment weighting (IPTW), a weight-based method for confounding adjustment. With these components established, we present the combined IPTW-IPCW estimator and its formulation. We also introduce Restricted Mean Survival Time (RMST) and Hazard Ratio (HR) as primary methods for evaluating treatment effect differences across survival curves.

### Problem Setup, Notation, and Assumptions

We discuss the problem of treatment effect estimation under the potential outcome framework in causal inference ^28,32^.

Consider a dataset with *n* independent and identically distributed patients, indexed by *i* ∈ ℐ = {1, …, *n*} Patient baseline covariates are denoted by **X**_*i*_ = (*X*_*i*1_, …, *X*_*ij*_) ∈ ℝ^*j*^ and the treatment variable is *Z*_*i*_ ∈{0, 1}.

Let 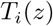 be the potential survival time and *C*_*i*_(*z*) be the potential censoring time if subject *i* had been assigned the treatment *z*. Then, *T*_*i*_ = *Z*_*i*_*T*_*i*_(1) + (1 *− Z*_*i*_)*T*_*i*_(0), and the observed follow-up time is 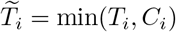 with the corresponding event indicator 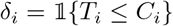 The observed dataset is thus 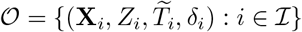.

Our primary goal is to estimate treatment effects in the context of time-to-event data subject to right censoring. We evaluate treatment effect using two complementary estimands:

1. The **Hazard Ratio (HR)** between the treatment-specific hazard functions:

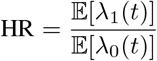

where *λ*_1_(*t*) is the hazard function under treatment, and *λ*_0_(*t*) is the hazard function under control.
2. The difference in **Restricted Mean Survival Time (RMST)** between the treatment groups. The RMST is defined as the expected survival time up to a specified time horizon, *τ*

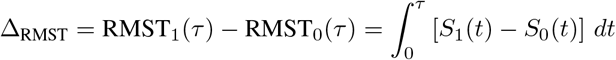

where *S*_1_(*t*) and *S*_0_(*t*) are the marginal survival functions for the treated and control groups, respectively.

We note that the HR formulation does not fully align with the standard potential outcomes framework, in which causal effects are defined in terms of contrasts between fixed potential outcomes at the individual level. In contrast, hazards represent instantaneous event rates and reflect population-level quantities ^9^. That is, hazard functions describe the rate of events over time among individuals with the same covariates, rather than a fixed risk for any one individual. Thus, the causal interpretation of hazard ratios should therefore be understood as a comparison of average instantaneous event rates across treatment groups rather than the average of individual-level causal effects.

To identify treatment effects in the presence of censoring, we rely on two sets of assumptions. The first set of assumptions, standard in causal inference, was introduced by Rosenbaum and Rubin ^27^ and allows us to estimate the causal effect of treatment in the absence of censoring. When these assumptions hold, we can interpret differences in observed survival outcomes as causal effects of treatment rather than biases due to confounding.

#### Assumption 1.

Stable Unit Treatment Value Assumption (SUTVA). *For every individual i:*

a. ***No interference***. *The potential survival time for subject i depends only on their own treatment, not on the treatments assigned to others; that is*,

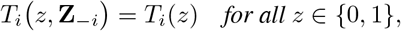

*where* **Z**_*−i*_ *denotes the treatment vector of all individuals except i*.
b. ***Consistency***. *The observed survival time in real data equals the potential outcome under the treatment actually received:*

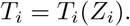

*Together, (a) and (b) comprise SUTVA; they rule out both interference across subjects and hidden versions of the treatment, ensuring each unit has a well-defined, unique potential outcome for every treatment level*.

#### Assumption 2.

Conditional Ignorability. *Treatment assignment is independent of potential survival times, conditional on observed baseline covariates:*

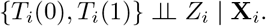

#### Assumption 3.

Positivity. *Each individual has a nonzero probability of receiving either treatment:*

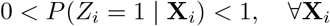

Assumptions 1-3 allow us to identify the causal effect of treatment on survival outcomes, provided that censoring does not introduce additional biases. To ensure that censoring does not obscure the identification of treatment effects, we introduce a second set of assumptions drawn from the causal survival analysis literature ^10,26,36^. The following assumptions are analogous assumptions 2 and 3, ensuring that censoring does not introduce selection bias.

#### Assumption 4.

Conditional Independence of Censoring. *Censoring is independent of the potential survival time, conditional on treatment assignment and baseline covariates:*

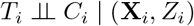

*i*.*e*., *after conditioning on* (**X**_*i*_, *Z*_*i*_), *the hazard of censoring carries no additional information about the future event time*.

Importantly, this conditional independence does not rule out the presence of marginal associations between survival and censoring. Such relationships can still arise in the marginal distribution due to shared dependence on covariates or treatment assignment.

#### Assumption 5.

Positivity of Censoring. *Each individual has a nonzero probability of remaining uncensored up to time t, conditional on covariates and treatment assignment:*

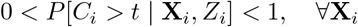

In this work, we focus on treatment effect estimation in scenarios where non-informative censoring assumption fails. We can relax that assumption to Assumption 4 and, together with Assumption 5, apply a data-driven model of the censoring process to construct inverse probability of censoring weights (IPCW) that remove the resulting informative censoring bias.

### Estimators

*Inverse Probability of Censoring Weighting (IPCW)* Inverse probability of censoring weighting (IPCW) provides a way to adjust for informative censoring by re-weighting observations based on their probability of remaining uncensored. IPCW works by assigning a weight to each subject at every observed time point *t*, inversely proportional to the estimated probability of remaining uncensored until time *t*. This probability is typically estimated using a separate model (often a Cox model or a pooled logistic regression), using censoring status as the dependent variable, to account for factors associated with censoring. When a subject is censored at time *t*_*c*_, those who remain at risk are given additional weight from *t*_*c*_ onward to compensate for the loss of information from the censored subject. Practically, IPCW weights vary over time per subject, updating each time the risk set changes.

IPCW corrects informative censoring only if the way subjects leave follow-up is adequately explained by observed data. In the present study, we only consider *baseline* covariates, so Assumptions 4 and 5 suffice. If time-varying covariates were available, one would need the stronger *sequential* ignorability of censoring assumption to hold ^26^.

The IPCW procedure can be summarized in the four steps below.

*Step 1* Fit a Cox PH model to estimate the hazard of censoring *λ*_*C*_(*t* | **X**_*i*_) conditional on covariates **X**_*i*_:

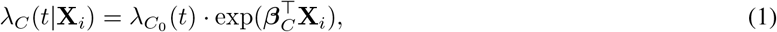

Where 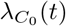 is the baseline hazard of censoring at time *t* and ***β***_*C*_ is the vector of model coefficients.

*Step 2* Estimate the probability of remaining uncensored up to each observed time point *t* for each subject.

Using the estimated hazard of censoring *λ*_*C*_(*t* | **X**_*i*_) from Step 1, we compute the subject-specific probability of remaining uncensored using the Cox model survival function:

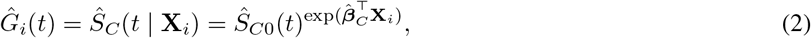

where *Ŝ*_*C*0_(*t*) is the estimated baseline survival function, corresponding to the probability of remaining uncensored up to time *t* for a subject with **X**_*i*_ = **0**. The baseline survival function is computed using the Breslow estimator:

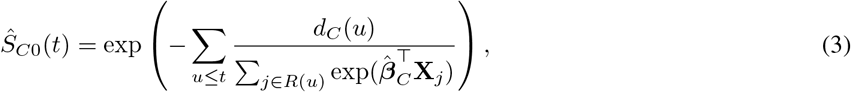

where *d*_*C*_(*u*) is the number of censoring events at time *u*, and *R*(*u*) is the risk set at time *u*.

*Step 3* Compute IPCW weights. For each subject *i* at time point *t*, a subject is at risk if 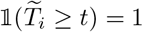 The inverse of the estimated probability of remaining uncensored up to time *t* is:

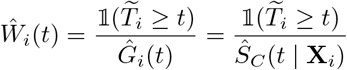

Stabilized IPCW weights can be computed by including the marginal (unconditional) probability of remaining uncensored *Ĝ*_0_(*t*) in the numerator:

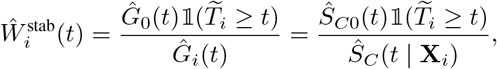

where *Ŝ*_*C*0_(*t*) = *Ĝ*_0_(*t*) is the baseline survival function estimated under the assumption that **X**_*i*_ = **0**.

*Step 4* Use IPCW weights in survival outcome models to estimate treatment effects (e.g., weighted Kaplan-Meier for survival probabilities, or weighted Cox proportional hazards model for hazard ratios).

The IPCW-adjusted Kaplan-Meier estimator is given by:

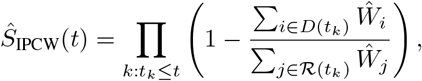

Where:

- *t*_*k*_ denotes the *k*-th ordered event time,
- *D*(*t*_*k*_) is the set of individuals who experience the event at time *t*_*k*_,
- *R*(*t*_*k*_) is the risk set just before *t*_*k*_,
- *Ŵ* _*i*_ is the IPCW weight for subject *i*.

Here, the numerator represents the weighted number of events at time *t*_*i*_, while the denominator represents the weighted number of individuals at risk. IPCW thus adjusts for the loss of information due to censoring.

To estimate treatment effects via a Cox proportional hazards model, IPCW weights can be incorporated into the partial likelihood function. The IPCW-adjusted partial likelihood is:

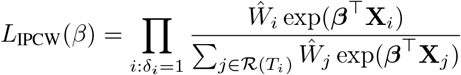

As with the unweighted model, estimates of ***β*** are obtained by maximizing the log of this weighted partial likelihood.

*Inverse Probability of Treatment Weighting (IPTW)* We briefly review Inverse Probability of Treatment Weighting (IPTW) as a standard method for confounding adjustment in observational studies here. IPTW is widely used to estimate causal effects by creating a pseudo-population in which treatment assignment is independent of observed confounders ^27^. By weighting individuals inversely to the probability of receiving the treatment they actually received, IPTW balances covariate distributions across treatment groups, enabling unbiased estimation of the treatment effect in the presence of confounding.

Since IPTW is well-established in the causal inference literature, we present it here only briefly as a review.

*Step 1* Fit a Propensity Score Model. We first estimate the propensity score *e*(**X**_*i*_), which is the conditional probability of receiving treatment given covariates:

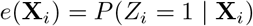

This is typically done using logistic regression. We note here that while IPCW weights are updated over time, each subject receives only a single IPTW weight, as treatment assignment is fixed at baseline.

*Step 2* Compute IPTW Weights. The weight for each subject *i* is given as the inverse of the estimated probability of receiving the treatment:

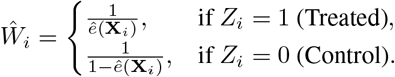

The stabilized IPTW weights for each individual *i* can be computed by taking the marginal probability of receiving the treatment and dividing it by the estimated probability of receiving the treatment:

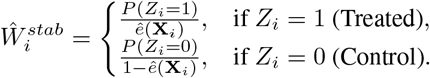

Here, *P* (*Z*_*i*_ = 1) and *P* (*Z*_*i*_ = 0) refer to the marginal treatment probabilities across the entire population.

*Step 3* Apply IPTW weights to outcome models. We apply the IPTW weights to estimated weighted Kaplan-Meier curves and weighted Cox Proportional Hazards models for survival probability and hazard ratio estimation, respectively.

For IPTW estimators to provide an unbiased correction for confounding, Assumptions 1-3 enumerated in section 2 (SUTVA, conditional ignorability, and positivity) must hold.

*Combining IPTW and IPCW* In observational studies with time-to-event outcomes, treatment effect estimation can be complicated by both confounding and informative censoring. To address both sources of bias simultaneously, we can construct a doubly weighted estimator by applying IPTW for confounding adjustment and IPCW for censoring correction ^3^.

The combined weight for each individual *i* is:

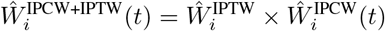

Here, 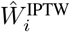 is constant over time, while 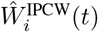 is a time-varying weight that adjusts for censoring. As a result, 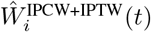 is also time-varying.

The identification of this combined estimator is shown in the appendix. Schaubel and Wei ^31^ demonstrated that this doubly weighted approach yields estimates that consistently recover the effect one would observe in a population where treatment was randomized and no censoring occurred.

## Simulation Study

To evaluate the performance of IPTW, IPCW, and their combination in correcting confounding and informative censoring, we conducted a simulation study. In all three simulations, we systematically varied the strength of informative censoring to assess the robustness and bias of each method under increasing levels of informative censoring bias. Simulation 1 featured non-differential informative censoring; simulation 2 introduced differential censoring; and simulation 3 incorporated differential censoring alongside a violation of the proportional hazards assumption.

We evaluated the performance of IPTW, IPCW, and their combination by comparing the following estimators:

- **Kaplan-Meier (KM) Estimator:** We assessed how accurately each method estimated survival probabilities over time, stratified by treatment group. Estimated survival curves were compared to those from a fully observed (uncensored) dataset to quantify bias introduced by censoring.
- **Restricted Mean Survival Time (RMST):** To evaluate absolute treatment benefit, we computed the difference in RMST between two treatment arms, up to a fixed time horizon *τ* = 500. Estimates were benchmarked against RMST values from the uncensored dataset.
- **Cox Proportional Hazards Model (Hazard Ratio (HR) Estimation):** We assessed each method’s ability to recover the true marginal hazard ratio specified during data generation.

We now describe the simulation setup in more detail.

### Simulation Design

*Step 1: Simulate covariate set*. We simulated a covariate matrix ***X***, which included a mix of continuous variables sampled from a normal distribution and binary variables sampled from a Bernoulli distribution.

*Step 2: Assign treatment status*. We assigned treatment status to each individual based on some subset of the covariates, denoted ***X***_*Z*_. We computed the probability of receiving treatment (the propensity score) as:

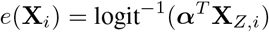

Here, ***α*** is the coefficient vector and **X**_*Z*,*i*_ denotes the treatment-relevant covariates for subject *i*. Treatment assignment *Z*_*i*_ was then sampled from a Bernoulli distribution *Z*_*i*_ = Bernoulli(*e*(**X**_*i*_))

*Step 3: Specify the ground-truth time-to-event model with a Weibull distribution*. For each individual *i*, the censoring time *T*_*i*_ was sampled from a Weibull distribution parametrized by shape parameter *k*_event_ and scale parameter *λ*_event_. Event times *T*_*i*_ were generated using inverse transform sampling as:

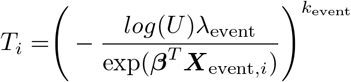

where *U*_*i*_ ~ Uniform(0, 1), ***β*** is the vector of log hazard ratios associated with covariates ***X***_event,*i*_, and the covariate set ***X***_event,*i*_ ⊆ ***X***_*i*_ is the subset of covariates influencing the event hazard. This subset may overlap with ***X***_*Z*,*i*_, the covariates affecting treatment assignment. This step was implemented using the R package simstudy ^11^.

This procedure yields a baseline hazard function:

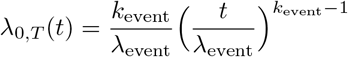

and individual-specific hazard functions conditional on covariates:

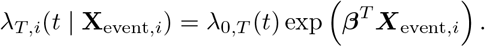

where *λ*_*T*,*i*_(*t* | ***X***_event,*i*_) is the hazard of the event for individual *i* at time *t*, and *λ*_0,*T*_ (*t*) is the baseline hazard function.

*Step 4: Specify the ground-truth time-to-censoring model with a Weibull distribution*. For each individual *i*, the censoring time *C*_*i*_ was sampled from a Weibull distribution parametrized by shape parameter *k*_censor_ and scale parameter *λ*_censor_. Censoring times *C*_*i*_ were generated using inverse transform sampling as:

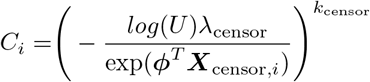

where *U*_*i*_ ~ Uniform(0, 1), ***ϕ*** is a vector of linear predictors that scales the impact of each covariate on the censoring hazard, and ***X***_censor,*i*_ ⊆ ***X***_*i*_ is the subset of covariates relevant for censoring. This subset may overlap with both ***X***_*Z*,*i*_ and ***X***_event,*i*_. This step was implemented using the R package simstudy ^11^.

This procedure yields a baseline *censoring* hazard function:

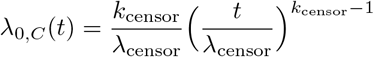

and individual-specific hazard functions conditional on covariates:

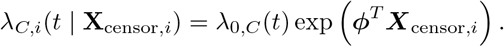

where *λ*_*C*,*i*_(*t* | ***X***_censor,*i*_) is the hazard of censoring for individual *i* at time *t*, and *λ*_0,*C*_(*t*) is the baseline hazard.

*Step 5: Sample event times and censoring times for each simulated individual in the dataset*. From the generated exponential distributions in steps 3 and 4, we sampled ground-truth event times *T*_*i*_ and ground-truth censoring times *C*_*i*_ for each individual. We computed the *observed* time for each individual, 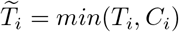 and the corresponding event indicator, 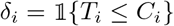.

*Step 6: Perform inference using simulated dataset*. With the simulated dataset 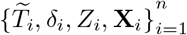 constructed from the previous steps, we estimated survival curves and treatment effects using weighted Kaplan-Meier estimators and Cox proportional hazards models.

To evaluate estimator performance, we compared estimates using each adjustment method (IPTW, IPCW, and combined IPCW+IPTW) against known ground-truth quantities derived from the full data-generating process:

- **Survival curves**: The ground truth survival function was computed using the uncensored event times *T*_*i*_ for all individuals, that is, event times assuming no censoring occurred. This provided the “true” survival function stratified by treatment group. To quantify discrepancies between estimated and true survival functions, we calculated the root mean squared error (RMSE). We first found the squared differences between the estimated and true survival probabilities across all time points in some pre-specified time analysis window (i.e., up to time *t* = *τ*) in both treatment groups, averaged across all computed squared differences, then took the square root. The RMSE served as a single summary measure of curve-level accuracy.
- **Restricted Mean Survival Time (RMST)**: The ground truth RMST was computed using the uncensored event times *T*_*i*_ for all individuals, in the same manner as the ground truth survival functions. Discrepancies between estimated and true RMST values were summarized as the RMST bias, defined as (estimate *−* truth).
- **Hazard ratio (HR)**: We simulated counterfactual event times, *T*_*i*_(1) and *T*_*i*_(0) for each individual. The true marginal hazard ratio was estimated by fitting a Cox PH model using the complete dataset of all potential outcomes (i.e., both factual and counterfactual event times), with no censoring. The model used treatment assignment as the independent variable and the true event time as the dependent variable. Discrepancies between estimated and true HR values were summarized as the HR bias, defined as (estimate *−* truth).

### Simulation Parameters

Our covariate set ***X*** consisted of four baseline variables: Conf1 (𝒩 (0, 0.5)), Conf2 (Bernoulli(0.5)), Cens1 (𝒩 (0, 0.5)), and Cens2 (Bernoulli(0.75)). Our covariate set had two confounders, Conf1 and Conf2, which were included in the treatment assignment model, i.e., {Conf1, Conf2} ⊆ ***X***_*Z*_. There were also two predictors for censoring, Cens1 and Cens2, which were included in the censoring model, i.e., {Cens1, Cens2} ⊆ ***X***_censor_. All four covariates, along with treatment assignment *Z*, influenced the event hazard, such that {Conf1, Conf2,} *Z*, Cens1, Cens2 ⊆ ***X***_event_.

The relationship among covariates, treatment, event, and censoring mechanisms is illustrated in Figure 2. We set *β*_*Z*_ = 0.7, corresponding to a *conditional* treatment effect size of 0.7 given the covariates. Each simulation was conducted with a sample size of *n* = 2000 and repeated 100 times.

**Figure 2.**
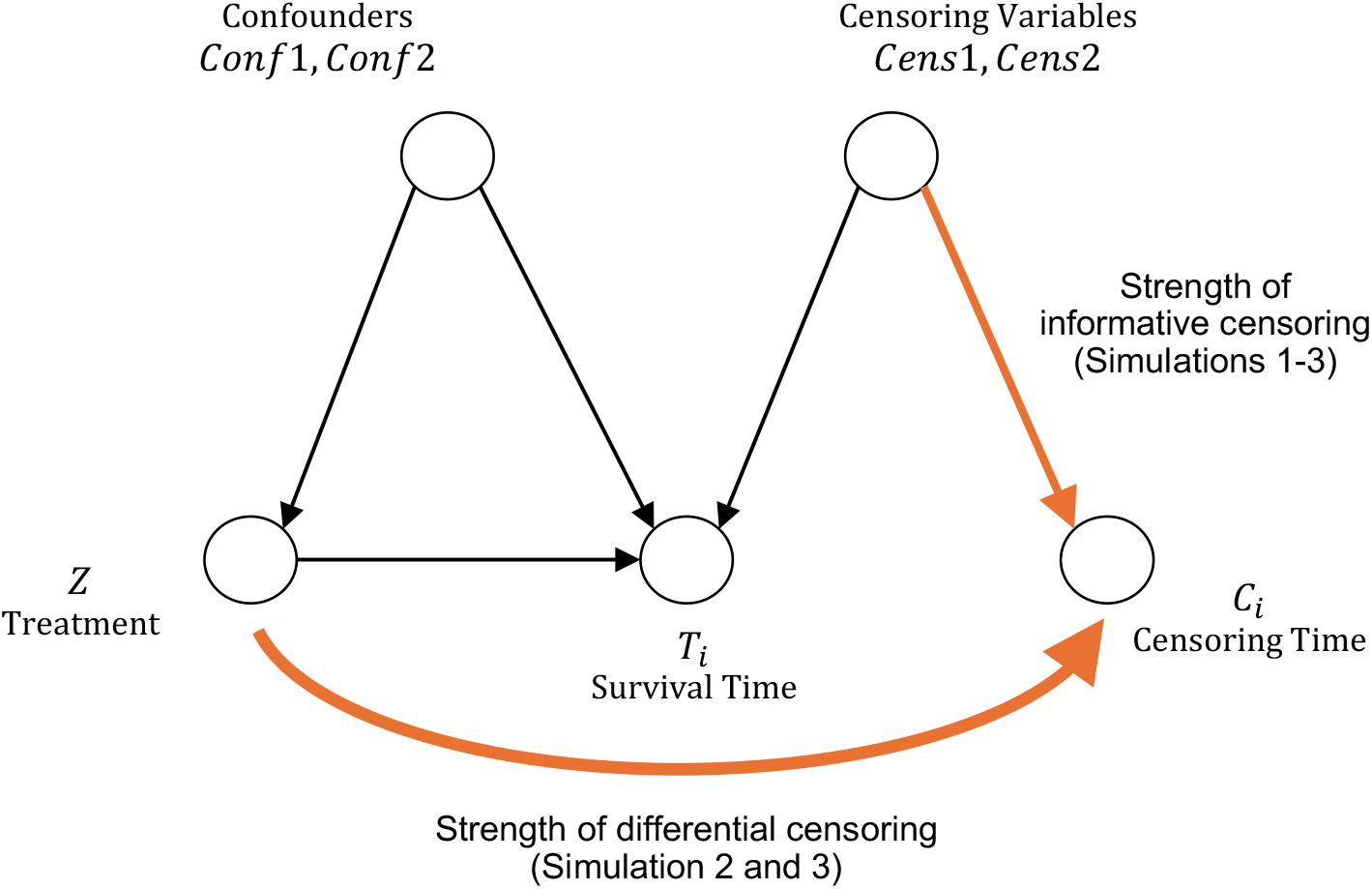
Simulation Setup. Bolded arrows denote what was varied in the simulation (informative censoring strength in all three simulations, and differential censoring in Simulations 2 and 3).

*Simulation 1: Varying censoring bias under non-differential informative censoring*. In Simulation 1, we varied the strength of informative censoring by modifying the parameter vector ***ϕ***, which governed the effect of Cens1 and Cens2 on the hazard of censoring. Increasing the magnitude of ***ϕ*** would lead to a stronger influence of Cens1 and Cens2 on the probability of being censored. Because these variables also affected the event outcome, larger magnitudes of ***ϕ*** induced greater informative censoring bias.

We evaluated four levels of informative censoring:

- **None: *ϕ*** = (*ϕ*_Cens1_ = 0, *ϕ*_Cens2_ = 0)
- **Weak: *ϕ*** = (0.8, 0.2)
- **Strong: *ϕ*** = (2, 0.8)
- **Weak Negative: *ϕ*** = (*−*0.8, *−*0.2)

For each value of ***ϕ***, the baseline hazard of censoring *λ*_0,*C*_(*t*) was adjusted to maintain an overall censoring rate of approximately 35%. Confounding bias was held constant at *α*_Conf1_ = 0.75 and *α*_Conf2_ = 0.25.

*Simulation 2: Varying censoring bias under differential informative censoring*. In Simulation 2, we introduced *differential* informative censoring by making the treatment variable *Z* associated with *both* the survival time and censoring time, i.e. {*Z*, Cens1, Cens2} ⊆ ***X***_censor_. We varied the strength of differential informative censoring as follows:

- **None: *ϕ*** = (*ϕ*_Z_ = 0, *ϕ*_Cens1_ = 0, *ϕ*_Cens2_ = 0)
- **Weak: *ϕ*** = (1.5, 0.8, 0.2)
- **Strong: *ϕ*** = (1.5, 2, 0.8)
- **Weak Negative: *ϕ*** = (*−*1.5, *−*0.8, *−*0.2)

The level of confounding and rate of censoring was held at the same level as Simulation 1.

*Simulation 3: Varying censoring bias under differential informative censoring and proportional hazards assumption violation*. In addition to varying the strength of informative censoring like Simulation 2, we generated data that violated the proportional hazards (PH) assumption by introducing a time-varying treatment effect.

Recall that the true time-to-event outcome was generated using a Weibull model, with hazard function:

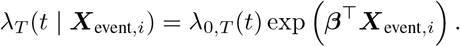

and that the scale parameter for the Weibull distribution was given by:

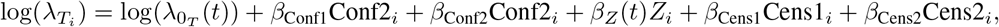

To introduce non-proportional, time-varying hazard ratios between the two treatment arms, we defined the treatment effect *β*_*Z*_(*t*) to be:

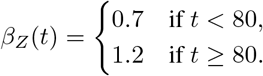

We used the same levels of confounding and rate of censoring as Simulations 1 and 2.

### Simulation Results

*Simulation 1: Varying censoring bias under non-differential informative censoring*. For survival curve estimation, IPCW and the combined method performed the best when there was informative censoring (Figures 4b-d). For RMST, IPTW-alone was generally sufficient. Although the combined method better captured the *survival curve*, both the IPTW method and combined methods captured the true RMST value consistently across varying levels of informative censoring (Table 1): this could be explained by the fact that although the IPTW-only survival curves were biased, both the treated and untreated curves were biased in the same direction (Figure 3), leading to an unbiased RMST difference between the treatment arms. For HR estimation (Table 1; Figure 4b), the combined method and IPTW-alone both captured the true marginal HR across different levels of informative censoring.

**Table 1.** Bias in RMST difference, hazard ratio (HR), and survival curves across different levels of informative censoring from **Simulation 1** (Varying censoring bias under non-differential informative censoring). RMST and HR bias is calculated as (estimate – truth); values closest to 0 indicate lower bias. Survival curve bias is summarized using the root mean squared error (RMSE), calculated as the square root of the average mean squared error between estimated and true survival probabilities, averaged across treatment and control arms and over time points from 0 to 500.

| Simulation scenario | Method | RMST bias (days) | HR bias | Survival RMSE |
| --- | --- | --- | --- | --- |
| No Informative Censoring | Unadjusted | -31.69 (-47.80, -15.58) | 0.42 (0.20, 0.64) | 0.038 |
|  | IPTW | -3.16 (-18.34, 12.02)<sup>†</sup> | 0.05 (-0.11, 0.22)<sup>†</sup> | 0.015 |
|  | IPCW | -31.59 (-47.67, -15.51) | 0.42 (0.20, 0.64) | 0.038 |
|  | IPTW + IPCW | -3.10 (-18.23, 12.04)<sup>†</sup> | 0.05 (-0.11, 0.22)<sup>†</sup> | 0.015 |
| Weak Informative Censoring | Unadjusted | -26.02 (-40.42, -11.62) | 0.45 (0.22, 0.67) | 0.041 |
|  | IPTW | -0.54 (-13.80, 12.72)<sup>†</sup> | 0.07 (-0.09, 0.24)<sup>†</sup> | 0.034 |
|  | IPCW | -31.96 (-49.11, -14.81) | 0.47 (0.23, 0.71) | 0.039 |
|  | IPTW + IPCW | -4.13 (-20.57, 12.32)<sup>†</sup> | 0.08 (-0.10, 0.26)<sup>†</sup> | 0.020 |
| Strong Informative Censoring | Unadjusted | -22.09 (-35.59, -8.60) | 0.48 (0.25, 0.71) | 0.064 |
|  | IPTW | 2.23 (-9.62, 14.08)<sup>†</sup> | 0.08 (-0.08, 0.25)<sup>†</sup> | 0.065 |
|  | IPCW | -33.58 (-61.07, -6.08) | 0.51 (0.14, 0.89) | 0.046 |
|  | IPTW + IPCW | -5.42 (-33.20, 22.37)<sup>†</sup> | 0.11 (-0.19, 0.41)<sup>†</sup> | 0.031 |
| Weak Informative Censoring (Negative) | Unadjusted | -40.90 (-57.55, -24.25) | 0.48 (0.25, 0.71) | 0.060 |
|  | IPTW | -7.87 (-24.59, 8.85)<sup>†</sup> | 0.07 (-0.10, 0.25)<sup>†</sup> | 0.033 |
|  | IPCW | -30.66 (-45.96, -15.35) | 0.39 (0.17, 0.60) | 0.037 |
|  | IPTW + IPCW | -2.26 (-17.24, 12.72)<sup>†</sup> | 0.03 (-0.13, 0.20)<sup>†</sup> | 0.016 |

<sup>†</sup> indicates the 95% confidence interval captures the true value.

**Figure 3.**
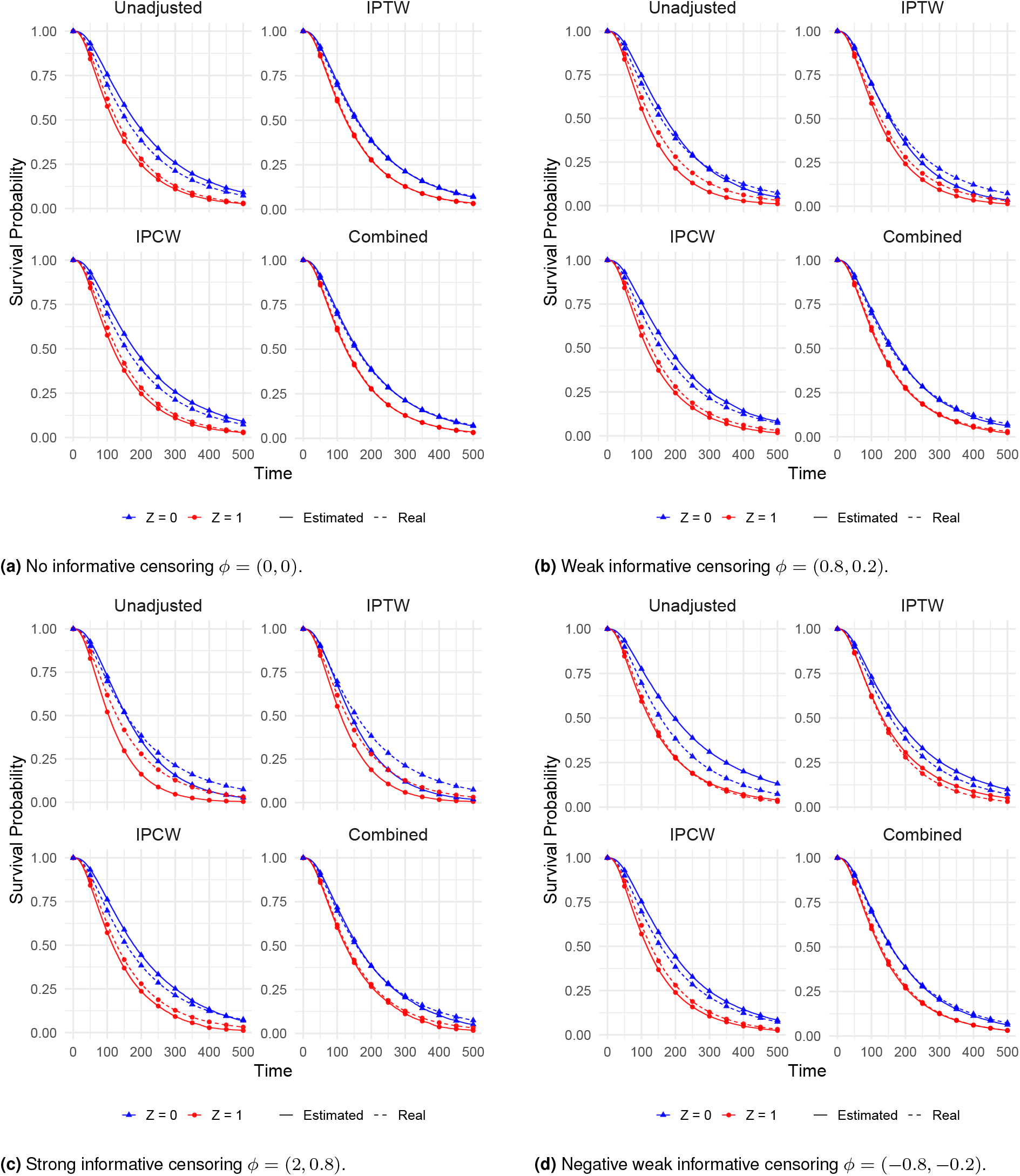
Treatment level survival curves when varying informative censoring strength in Simulation 1 (Varying censoring bias under non-differential informative censoring). *ϕ* values denote the relationships of the covariates “Cens1” and “Cens2” to censoring, respectively. Dotted lines denote the ground-truth survival curves, i.e. event times in the absence of censoring.

**Figure 4.**
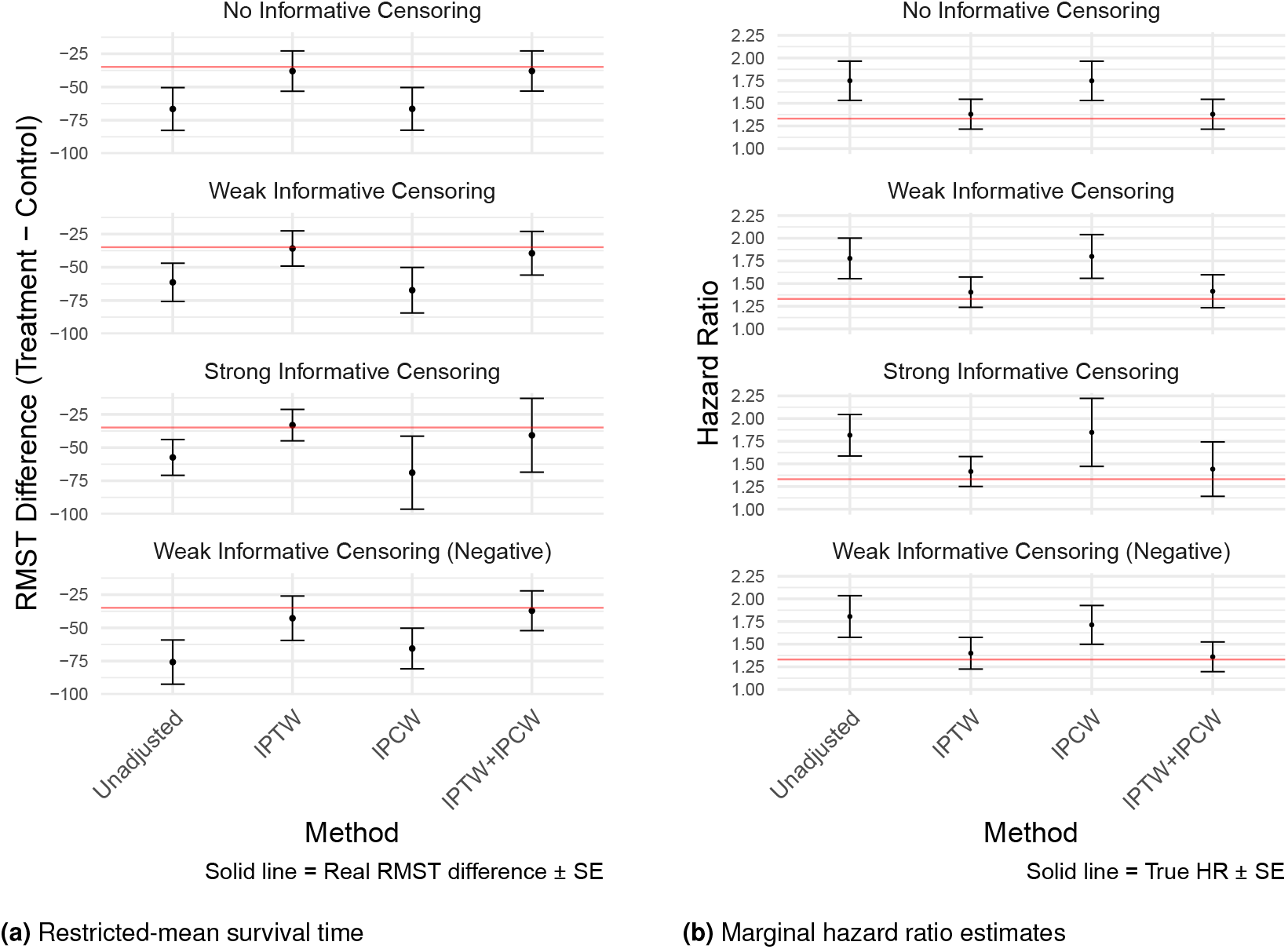
Treatment Effect Estimation for Simulation 1 (Varying censoring bias under non-differential informative censoring): comparison of four estimators for (a) RMST difference at *τ* = 500 and (b) marginal hazard ratio estimates when varying the strength of informative censoring, *ϕ*.

*Simulation 2: Varying censoring bias under differential informative censoring*. For survival curve estimation, the combined method performed the best across different levels of informative censoring (Figure 5). For RMST (Table 2; Figure 6a) and HR estimation (Table 2; Figure 6b), the combined method was the only estimator that consistently recovered the ground truth under informative censoring. Notably, the results from both IPCW and the combined methods had larger standard errors in the presence of strong informative censoring, which is possibly due to the spread of the IPCW weights as more subjects get censored over time. This larger standard error was seen for both RMST and HR estimation.

**Table 2.**
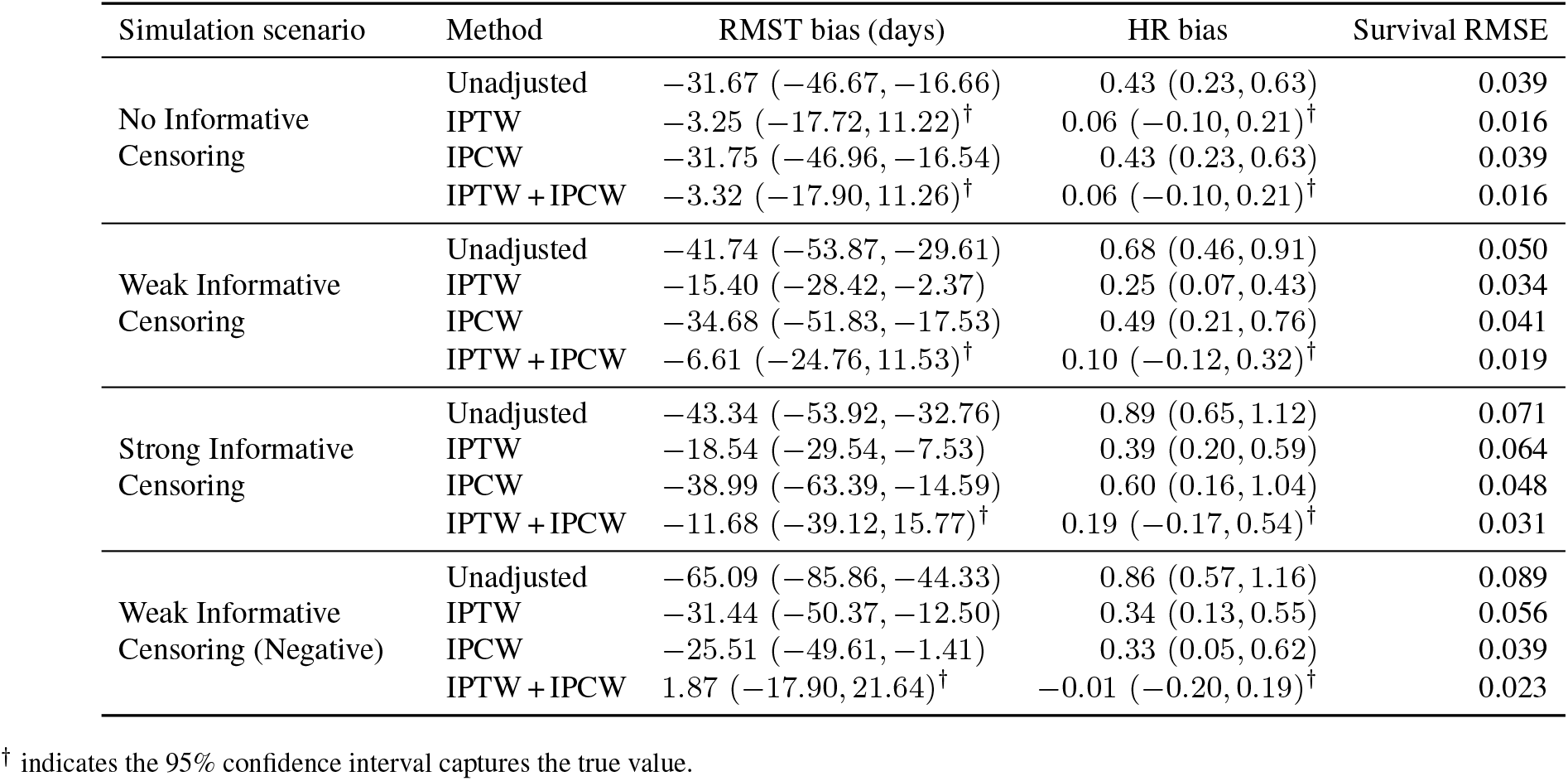
Bias in RMST difference, hazard ratio (HR), and survival curves across different levels of informative censoring from **Simulation 2** (Varying censoring bias under differential informative censoring). RMST and HR bias is calculated as (estimate – truth); values closest to 0 indicate lower bias. Survival curve bias is summarized using the root mean squared error (RMSE), calculated as the square root of the average mean squared error between estimated and true survival probabilities, averaged across treatment and control arms and over time points from 0 to 500.

| Simulation scenario | Method | RMST bias (days) | HR bias | Survival RMSE |
| --- | --- | --- | --- | --- |
| No Informative Censoring | Unadjusted | -31.67 (-46.67, -16.66) | 0.43 (0.23, 0.63) | 0.039 |
|  | IPTW | -3.25 (-17.72, 11.22)<sup>†</sup> | 0.06 (-0.10, 0.21)<sup>†</sup> | 0.016 |
|  | IPCW | -31.75 (-46.96, -16.54) | 0.43 (0.23, 0.63) | 0.039 |
|  | IPTW + IPCW | -3.32 (-17.90, 11.26)<sup>†</sup> | 0.06 (-0.10, 0.21)<sup>†</sup> | 0.016 |
| Weak Informative Censoring | Unadjusted | -41.74 (-53.87, -29.61) | 0.68 (0.46, 0.91) | 0.050 |
|  | IPTW | -15.40 (-28.42, -2.37) | 0.25 (0.07, 0.43) | 0.034 |
|  | IPCW | -34.68 (-51.83, -17.53) | 0.49 (0.21, 0.76) | 0.041 |
|  | IPTW + IPCW | -6.61 (-24.76, 11.53)<sup>†</sup> | 0.10 (-0.12, 0.32)<sup>†</sup> | 0.019 |
| Strong Informative Censoring | Unadjusted | -43.34 (-53.92, -32.76) | 0.89 (0.65, 1.12) | 0.071 |
|  | IPTW | -18.54 (-29.54, -7.53) | 0.39 (0.20, 0.59) | 0.064 |
|  | IPCW | -38.99 (-63.39, -14.59) | 0.60 (0.16, 1.04) | 0.048 |
|  | IPTW + IPCW | -11.68 (-39.12, 15.77)<sup>†</sup> | 0.19 (-0.17, 0.54)<sup>†</sup> | 0.031 |
| Weak Informative Censoring (Negative) | Unadjusted | -65.09 (-85.86, -44.33) | 0.86 (0.57, 1.16) | 0.089 |
|  | IPTW | -31.44 (-50.37, -12.50) | 0.34 (0.13, 0.55) | 0.056 |
|  | IPCW | -25.51 (-49.61, -1.41) | 0.33 (0.05, 0.62) | 0.039 |
|  | IPTW + IPCW | 1.87 (-17.90, 21.64)<sup>†</sup> | -0.01 (-0.20, 0.19)<sup>†</sup> | 0.023 |

<sup>†</sup> indicates the 95% confidence interval captures the true value.

**Figure 5.**
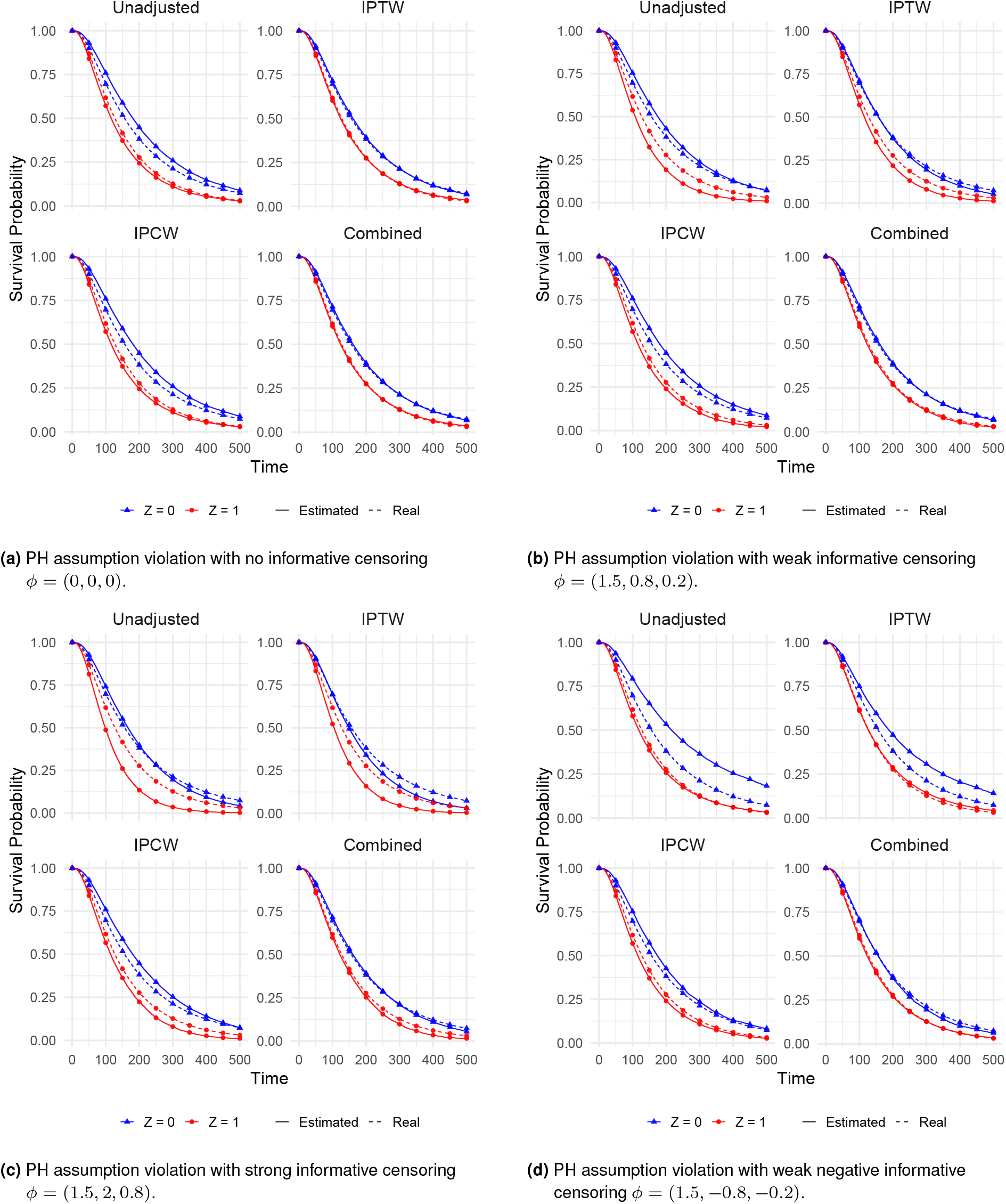
Treatment level survival curves when varying *differential* informative censoring strength in Simulation 2 (Varying censoring bias under differential informative censoring). *ϕ* values denote the relationships of the covariates “Z” (treatment), “Cens1”, and “Cens2” to censoring, respectively. Dotted lines denote the ground-truth survival curves, i.e. event times in the absence of censoring.

**Figure 6.**
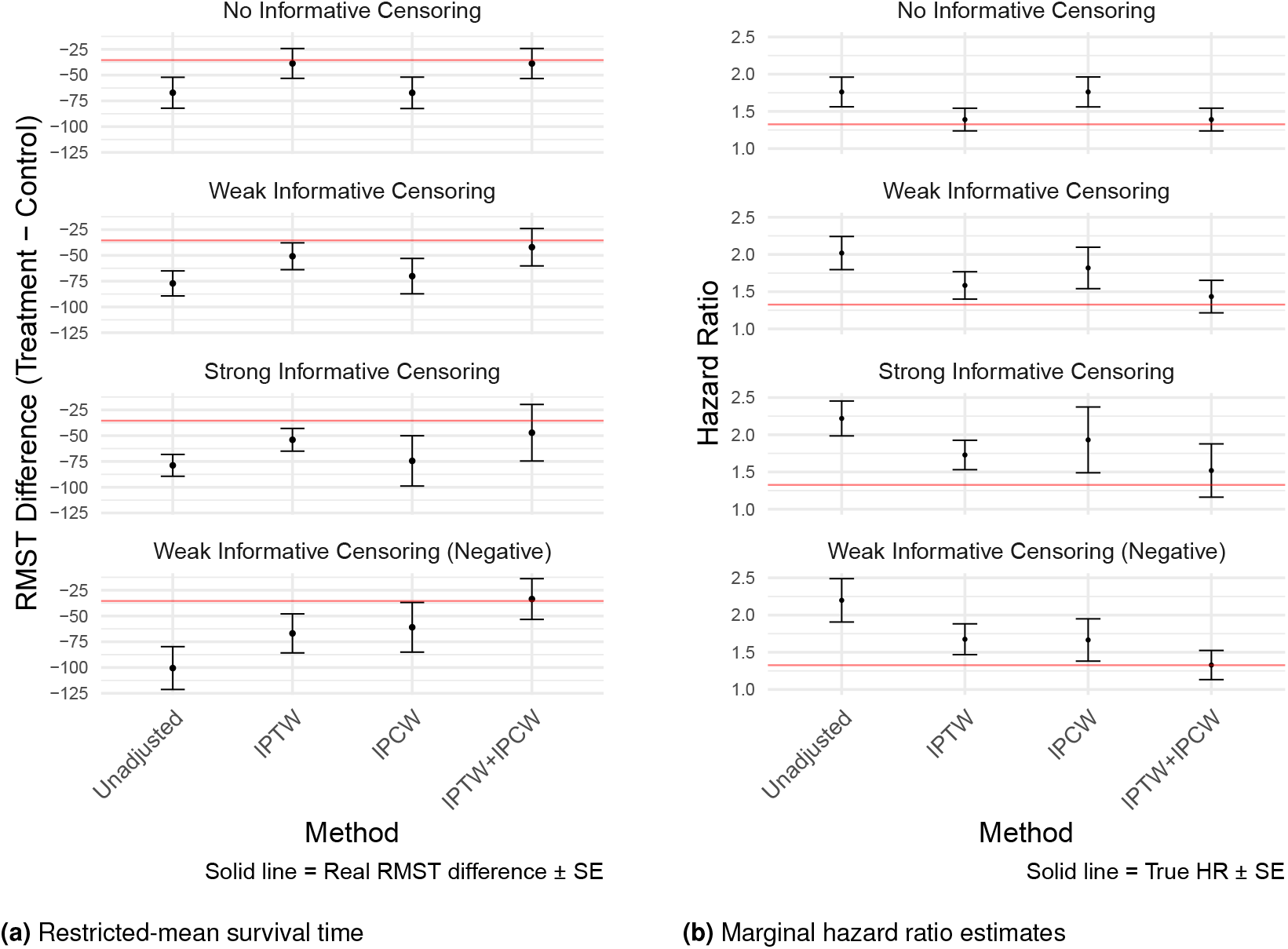
Treatment Effect Estimation for Simulation 2 (Varying censoring bias under differential informative censoring): comparison of four estimators for (a) RMST difference at *τ* = 500 and (b) marginal hazard ratio estimates when varying the strength of informative censoring, *ϕ*.

*Simulation 3: Varying censoring bias under differential informative censoring and proportional hazards assumption violation*. When the PH assumption was violated in the presence of differential informative censoring, the results were similar to the case where there was no PH violation: the combined method performed the best for survival curve estimation (Figure 7). The combined method was also the only method to consistently recover the true RMST (Table 3; Figure 8a) and true HR (Table 3; Figure 8b).

**Table 3.** Bias in RMST difference, hazard ratio (HR), and survival curves across different levels of informative censoring from **Simulation 3** (Varying censoring bias under differential informative censoring and proportional hazards assumption violation). RMST and HR bias is calculated as (estimate – truth); values closest to 0 indicate lower bias. Survival curve bias is summarized using the root mean squared error (RMSE), calculated as the square root of the average mean squared error between estimated and true survival probabilities, averaged across treatment and control arms and over time points from 0 to 500.

| Simulation scenario | Method | RMST bias (days) | HR bias | Survival RMSE |
| --- | --- | --- | --- | --- |
| No Informative Censoring | Unadjusted | -30.04 (-43.74, -16.33) | 0.49 (0.26, 0.72) | 0.038 |
|  | IPTW | -2.88 (-16.31, 10.55)<sup>†</sup> | 0.05 (-0.12, 0.23)<sup>†</sup> | 0.015 |
|  | IPCW | -30.10 (-44.12, -16.07) | 0.49 (0.26, 0.72) | 0.038 |
|  | IPTW + IPCW | -2.94 (-16.59, 10.71)<sup>†</sup> | 0.05 (-0.12, 0.23)<sup>†</sup> | 0.015 |
| Weak Informative Censoring | Unadjusted | -36.19 (-47.91, -24.47) | 0.77 (0.49, 1.04) | 0.044 |
|  | IPTW | -10.72 (-23.00, 1.56)<sup>†</sup> | 0.26 (0.03, 0.48) | 0.027 |
|  | IPCW | -31.70 (-46.49, -16.91) | 0.54 (0.23, 0.86) | 0.039 |
|  | IPTW + IPCW | -4.82 (-20.20, 10.56)<sup>†</sup> | 0.09 (-0.15, 0.34)<sup>†</sup> | 0.016 |
| Strong Informative Censoring | Unadjusted | -37.08 (-47.35, -26.81) | 1.02 (0.72, 1.31) | 0.060 |
|  | IPTW | -12.95 (-23.60, -2.29) | 0.44 (0.19, 0.68) | 0.052 |
|  | IPCW | -36.29 (-54.62, -17.96) | 0.71 (0.25, 1.18) | 0.044 |
|  | IPTW + IPCW | -10.16 (-29.98, 9.66)<sup>†</sup> | 0.22 (-0.15, 0.59)<sup>†</sup> | 0.025 |
| Weak Informative Censoring (Negative) | Unadjusted | -58.57 (-77.81, -39.32) | 0.90 (0.57, 1.22) | 0.080 |
|  | IPTW | -27.16 (-44.74, -9.58) | 0.33 (0.09, 0.56) | 0.047 |
|  | IPCW | -23.71 (-42.94, -4.48) | 0.39 (0.11, 0.68) | 0.035 |
|  | IPTW + IPCW | 2.27 (-14.14, 18.68)<sup>†</sup> | -0.01 (-0.21, 0.19)<sup>†</sup> | 0.019 |

<sup>†</sup> indicates the 95% confidence interval captures the true value.

**Figure 7.**
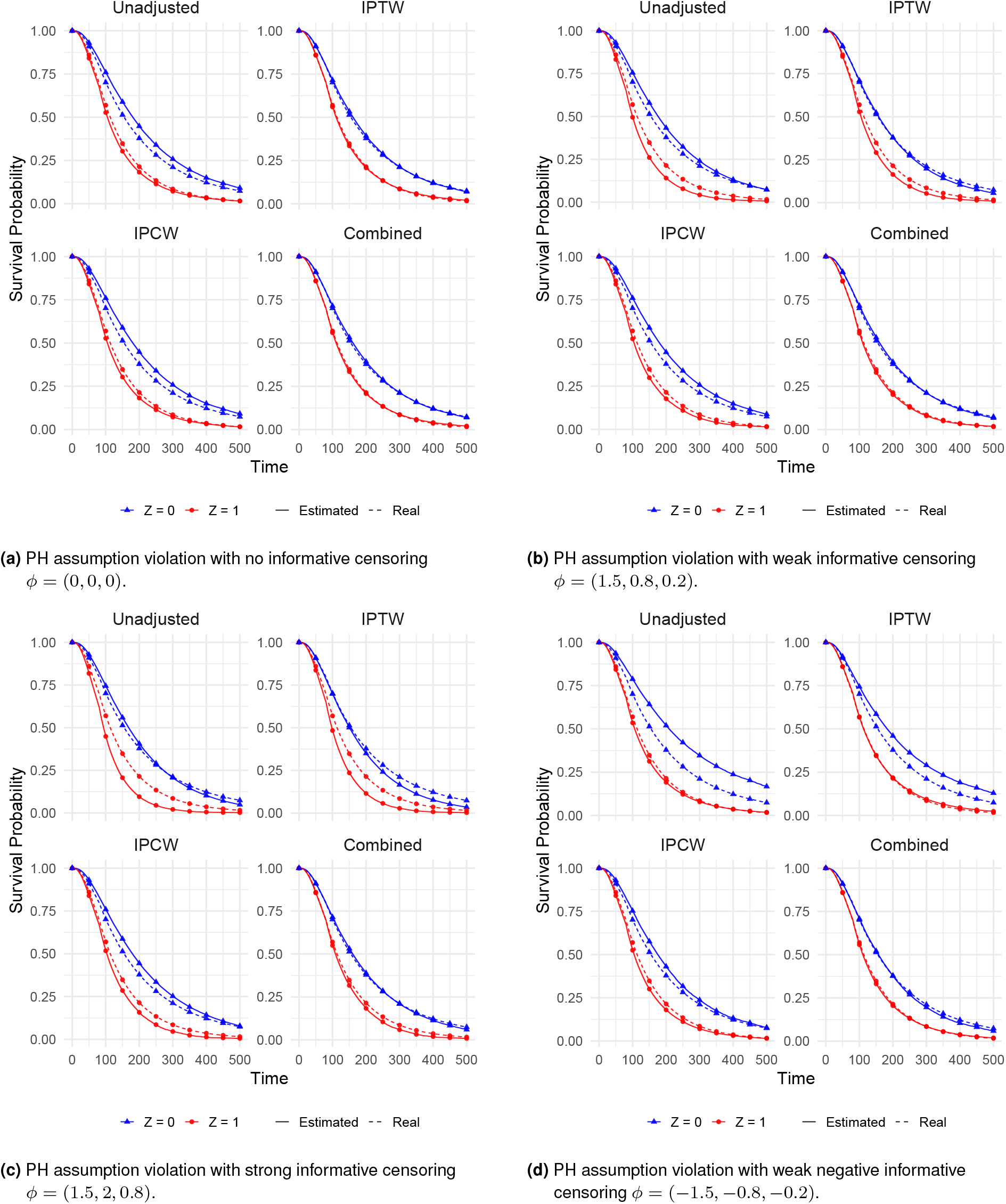
Treatment level survival curves when varying *differential* informative censoring strength and proportional hazards assumption violation in Simulation 3 ((Varying censoring bias under differential informative censoring and proportional hazards assumption violation). *ϕ* values denote the relationships of the covariates “Z” (treatment), “Cens1”, and “Cens2” to censoring, respectively. Dotted lines denote the ground-truth survival curves, i.e. event times in the absence of censoring.

**Figure 8.**
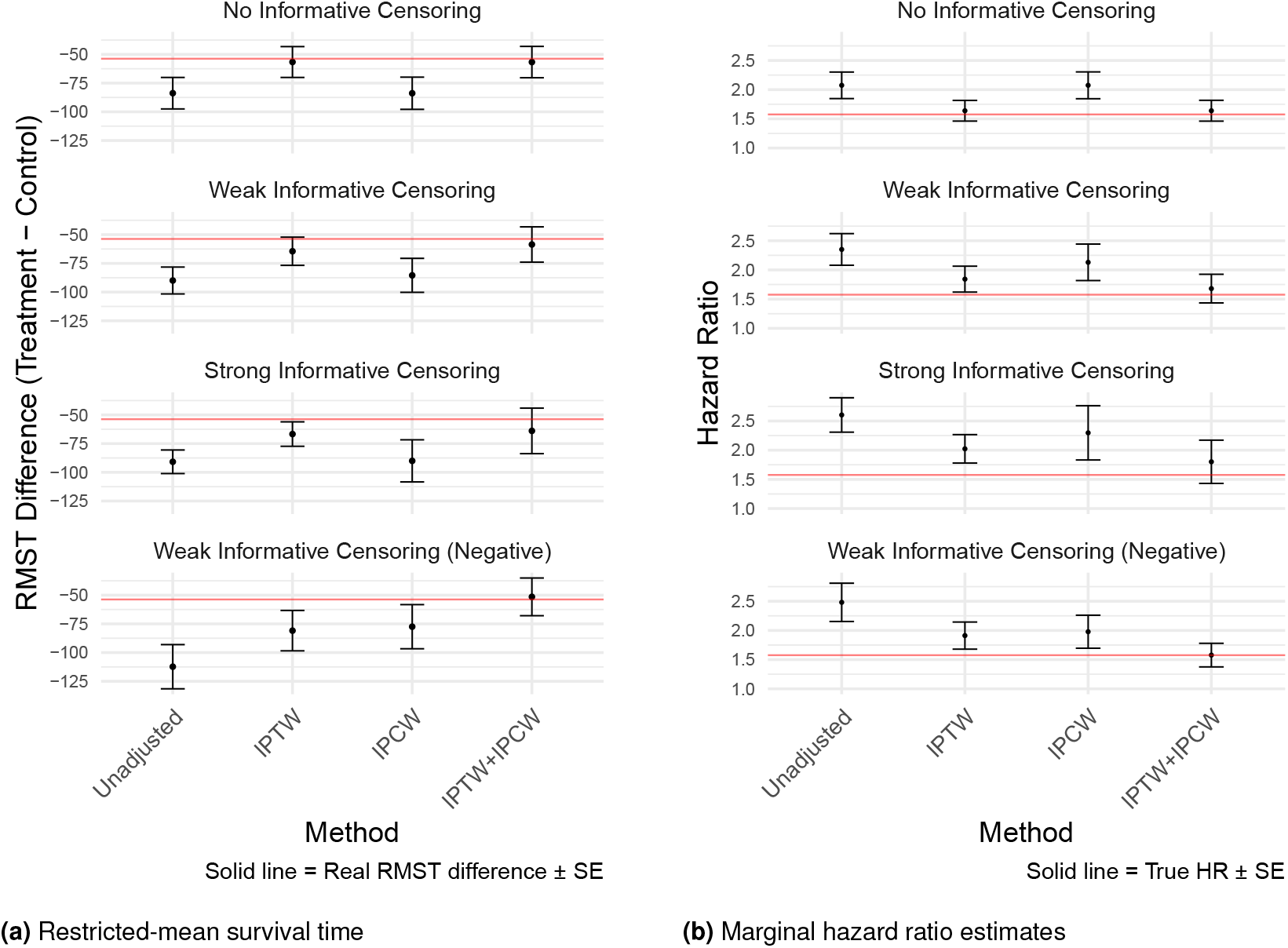
Treatment Effect Estimation for Simulation 3 (Varying censoring bias under differential informative censoring and proportional hazards assumption violation): comparison of four estimators for (a) RMST difference at *τ* = 500 and (b) marginal hazard ratio estimates when varying the strength of informative censoring, *ϕ*.

## Real World Study

We compared the effects of two classes of anti-hypertensive drugs (ACE inhibitors and thiazide/thiazide-like diuretics) on one clinical outcome, acute myocardial infarction (AMI), with a retrospective, observational, comparative new-user cohort design.

We used a single electronic health records database from Columbia University Medical Center (CUMC). We had a cohort size of 41,996 and a total of 49,104 covariates. We estimated the comparative treatment effect with Cox proportional hazards models and used 4 different weighting methodologies: (1) Unadjusted (no confounding or censoring adjustment), IPTW alone (confounding adjustment), IPCW alone (censoring adjustment) and the combined IPTW+IPCW weights (both confounding and censoring adjustment).

IPTW weights were calculated from the 49,104 covariates with Large Scale Propensity Scores (LSPS) ^40^, and IPCW weights were calculated using a L1-regularized Cox PH Model. Because updating the IPCW weights every time the risk set changed would result in 6,205 different refittings of the Cox censoring model (once per change in the risk set; whenever a patient was censored or had the event), we opted to update IPCW weights once per 60 day interval.

For each weighting method, we estimated the hazards ratio, the survival curves, and the difference in RMST between the two treatments. Standard errors of hazard ratios were estimated through a robust sandwich estimator ^20^, and standard errors of RMST differences were estimated with bootstrapping. We then compared HR estimates with results from a previously published large-scale meta-analysis with the same study design, involving 20 million patients across 9 databases and 4 countries ^33^ (LEGEND-HTN study). The 2019 study used propensity scores calculated from LSPS, but used 1:1 matching instead of IPTW weights. While we also calculated survival curves and RMST, there is no external benchmark for comparison, so we simply reported the results.

### Results

The survival curves (Figure 9) and RMST (Figure 10) estimates showed that the unadjusted and IPCW curves performed similarly, while IPTW and the combined curves performed similarly. The unadjusted analysis suggested a substantial 27-day gain in restricted mean survival time (RMST) for the thiazide group (95% CI 22-32 days). Censoring adjustment alone (IPCW) did little to change this estimate (26.8 days, 22-32 days). In contrast, weighting for treatment confounding with IPTW sharply attenuated the RMST difference to 3.3 days (−1.0-7.5 days), bringing the estimate closer to the null. Adding censoring weights on top of IPTW produced a similar result (2.6 days, −1.5-6.7 days).

**Figure 9.**
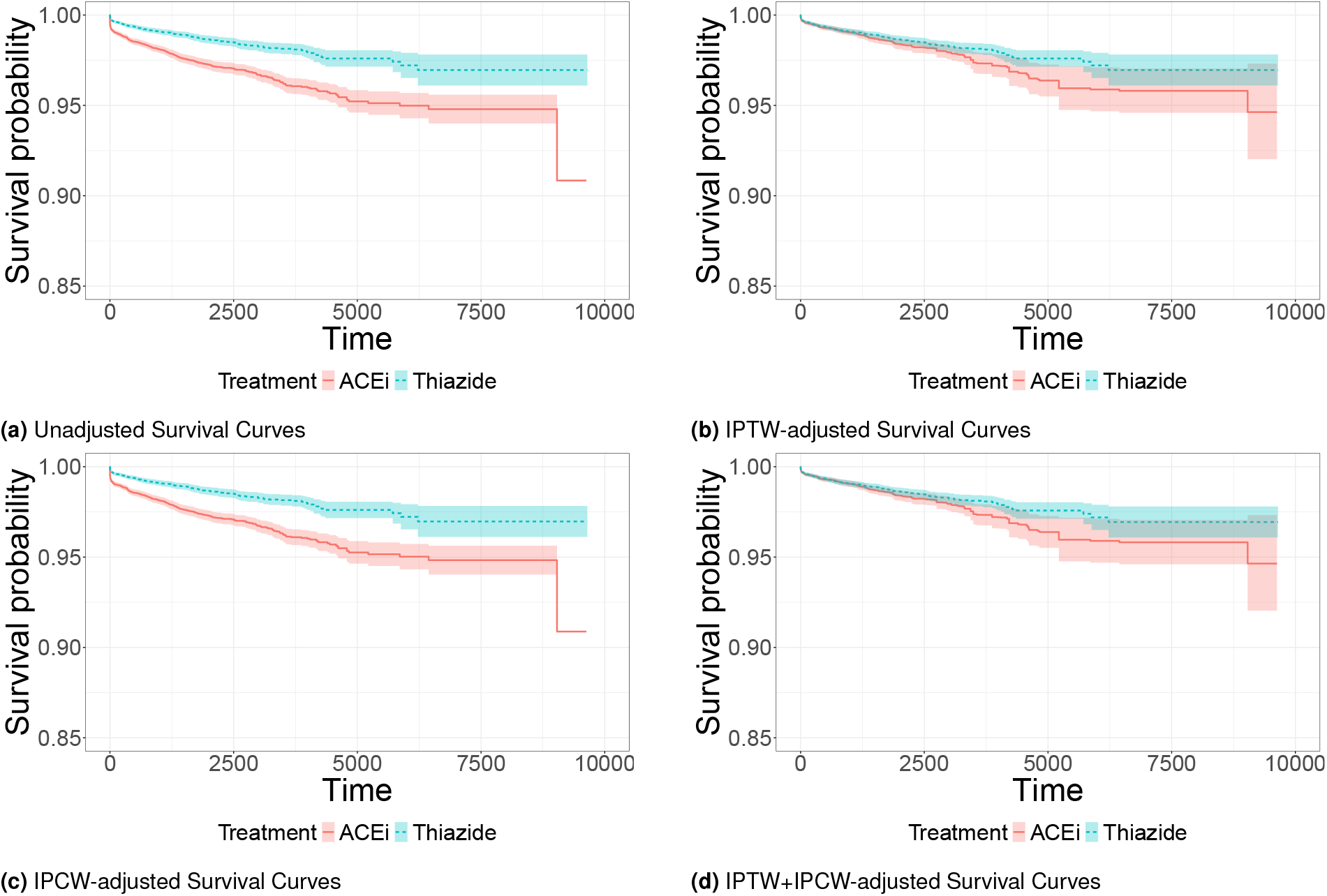
Unadjusted survival curves (a) did not use any weights, while survival curves are weighted by (b) IPTW, (c) IPCW, and (d) combining IPTW+IPCW. The two treatment groups were thiazide diuretics (dotted line) and ACE inhibitors (solid line). Shaded areas denote the 95% confidence interval.

**Figure 10.**
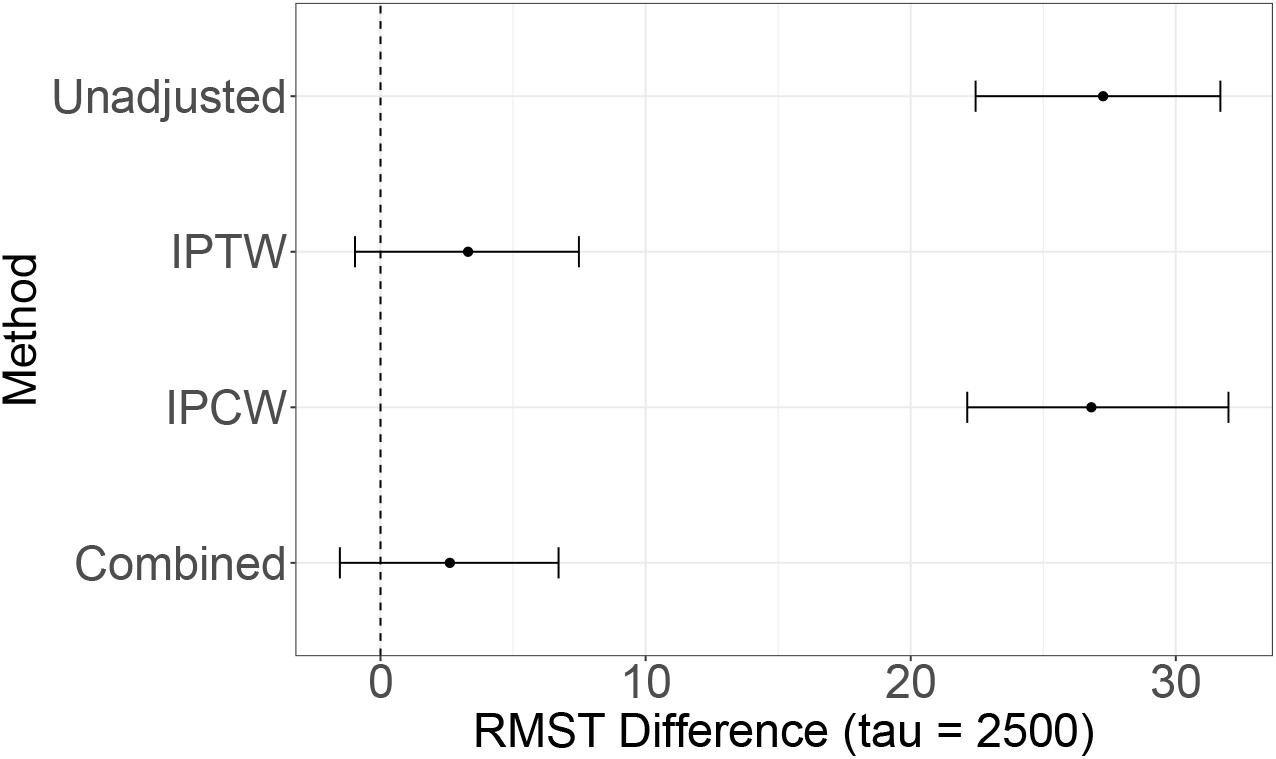
RMST differences in our real world study, comparing thiazide diuretics to ACE inhibitors for the risk of AMI. A RMST difference > 0 means that the outcome is more favorable for thiazide diuretics. The dotted vertical line denotes the point at which there is no difference in survival time between the two treatments. Standard errors were calculated through bootstrap (n=200).

For HR estimation (Figure 11), the unadjusted estimate underestimated the published benchmark (HR = 0.48, 95% CI 0.41–0.56 vs. 0.84, 0.75–0.95 in the LEGEND-HTN meta-analysis). Applying IPTW brought the hazard ratio much closer to the benchmark (HR = 0.81, 0.66–0.99). Adding censoring weights (combined IPTW + IPCW) yielded a slightly higher point estimate (HR = 0.92, 0.73–1.14), but the two approaches were not statistically significantlly different given their overlapping confidence intervals. In contrast, IPCW alone failed to improve on the unadjusted analysis, producing an HR of 0.46 (0.38–0.55).

**Figure 11.**
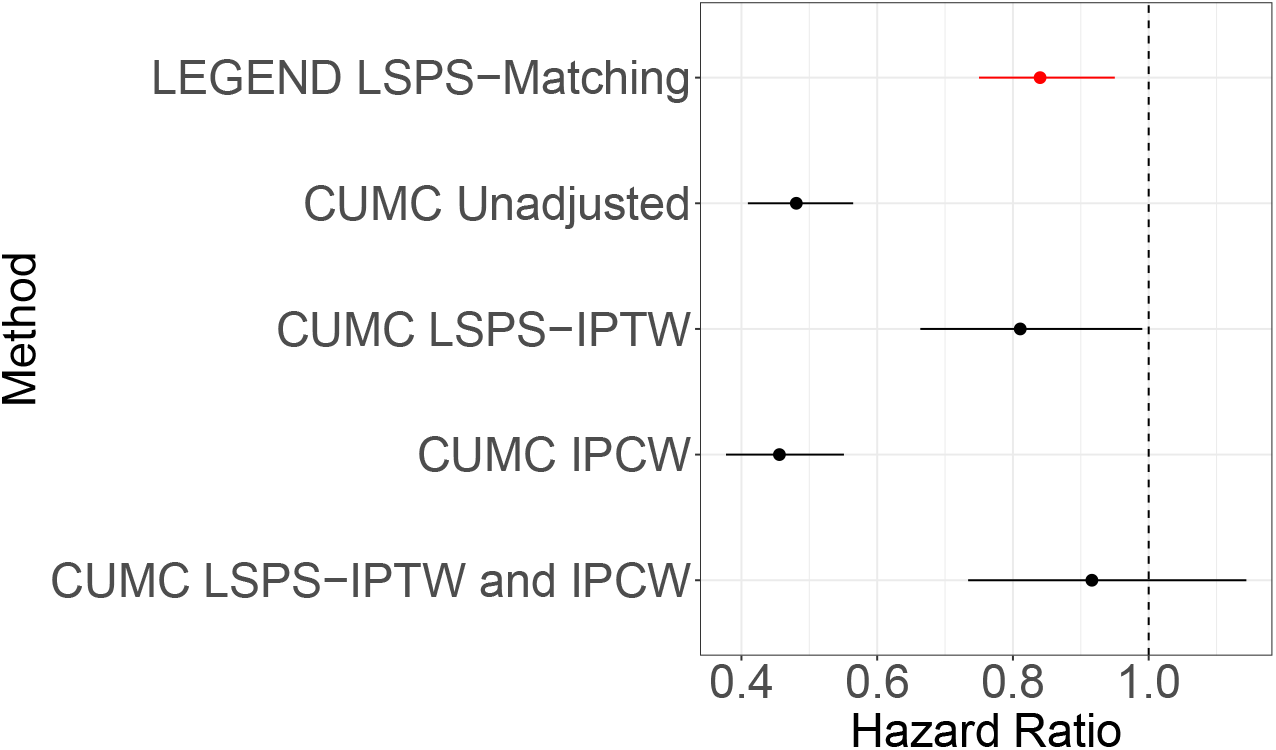
Hazard ratio estimates of our real world study, comparing thiazide diuretics to ACE inhibitors for the risk of AMI. A HR < 1 means that the outcome is more favorable for thiazide diuretics. The first line denotes the meta-analysis study (LEGEND-HTN) from Suchard et. al ^33^. All other estimates are from a single EHR database (“CUMC”). The dotted vertical line denotes the point at which there is no difference in hazard ratio between the two treatments. Standard errors were estimated with a robust sandwich-type variance estimator proposed by Lin and Wei ^20^.

## Discussion

This study evaluated the performance of IPTW, IPCW, and their combination in the presence of confounding and informative censoring. We assessed these weighting methods on survival curve, RMST, and HR estimation in simulation and real-world data settings.

Our simulations showed that IPCW and the combined method consistently produced survival curves closer to the ground-truth, while the addition of IPCW only improved upon treatment effect estimation (RMST and HR) in the presence of *differential* informative censoring. This is likely explained by the fact that, though non-differential informative censoring biases survival curves, both the treated and untreated curves had biases in the same direction with similar magnitudes (Figure 3), ultimately leading to a unbiased treatment effect estimate (either a difference or a ratio between the untreated and treated hazards or RMST).

In our real-world study, IPCW adjustment did not improve HR estimates relative to an unadjusted analysis when compared against the published large-scale meta-analysis. This could be due to the fact that the IPCW weights were updated too infrequently (once every 60 days, instead of every time the risk set changed). Another possible explanation could be that the primary source of bias in the LEGEND-HTN study was confounding, and that the informative censoring was absent or weak. The results from the real-world study showed that the IPCW estimates closely followed that of the unadjusted RMST, HR, and survival curve estimates, while the IPTW estimates closely followed that of the combined IPTW+IPCW RMST, HR, and survival curve estimates. This was seen as well in all 3 simulations in the case where there was no informative censoring.

Our findings suggest that the addition of IPCW is valuable for (1) survival curve estimation in the presence of informative censoring, and (2) treatment effect estimation in the presence of differential informative censoring. Practically, this suggests that if the primary goal is to estimate survival probabilities, or if it is known that there is presence of differential informative censoring, the doubly weighted method may yield more accurate results than IPTW alone. We note, however, that determining whether informative censoring is “strong” or “differential” in the real-world data setting is difficult: because we only observe *either* the censoring time or the event time for each individual, never both, the extent of informative censoring is unidentifiable. One potential strategy to assess the value of IPCW is to fit separate models for treatment assignment and censoring, then examine whether any covariates are predictive of censoring but not included in the treatment model. If such variables also affect the outcome, their omission from the confounding adjustment may lead to residual bias that IPTW cannot correct, whereas IPCW may help mitigate it. We leave the development of this diagnostic approach as a future direction.

### Limitations and Future Directions

There are some limitations of our study. First, this study focused on baseline confounders and censoring mechanisms; however, in clinical practice, time-dependent confounding and censoring are common. Second, while we compared our real-world study with an external benchmark ^33^, it is important to recognize that the meta-analysis itself is not necessarily the ground truth. In a follow-up study also comparing thiazide diuretics and ACE inhibitors for the risk of AMI, the study design was modified to use a stricter definition of antihypertensive monotherapy to mitigate bias ^1^. Though the results of this follow-up study do not differ substantially from the original meta-analysis, the risk of AMI shifted towards the null and was no longer significant. This shift towards the null was also seen in our IPTW+IPCW result. While the 2024 follow-up study and the study presented here attempted to correct bias from different sources, the fact that the IPTW+IPCW HR was closer to that of the 2024 follow-up study (than to the meta-analysis) suggests that there may exist some residual bias in the large-scale meta analysis. In addition, differences in study populations, data sources, and methodological choices (e.g., 1:1 propensity score matching in the meta-analysis vs. IPTW in our study) could also contribute to discrepancies. It is also possible that the IPTW estimate performed the “best” out of all four weighting methods when compared to the published meta-analysis because the meta-analysis itself did not adjust for censoring. However, we note that the meta-analysis might provide a better estimate even in the absence of censoring adjustment, because it used several large claims databases, which might capture more comprehensive observation periods than electronic health records ^2^. Given the lack of a true experimental gold standard, this comparison represents the best available approach to assess the plausibility of the hazards ratio estimates. Finally, our real-world study evaluated a treatment-outcome pair where informative censoring may not have played a major role, particularly after adjusting for confounders. It is possible that the role of IPCW would be more pronounced in scenarios where censoring is strongly informative and differential, and not fully explained by observed covariates. Future research could explore alternative treatment-outcome pairs where censoring is more likely to introduce bias in order to better understand the practical necessity of IPCW.

### Conclusions

This study underscores the importance of evaluating whether censoring meaningfully biases treatment effect estimation when deciding whether to apply censoring adjustment methods. The findings in our real-world study suggests that when treatment assignment is primarily driven by patient characteristics, large-scale confounder adjustment (e.g., using large-scale propensity scores) is often sufficient to remove most biases, including those indirectly related to censoring. In such cases, explicit censoring adjustment may not be necessary. However, in the presence of informative censoring (whether or not it is differential), IPCW improves survival curve estimation bias. Furthermore, if the informative censoring is also differential, the combination of IPCW and IPTW may be necessary to get unbiased treatment effect (RMST, HR) estimates.

## Data Availability

The data underlying this article cannot be shared publicly due to privacy of patient health information.

## Appendix

### Causal Identification of RMST Difference Using IPTW and IPCW

We now formally state and prove the identification result for the RMST difference

**Assumptions:**

**A1 Consistency**: *T* = *T* (*z*) when *Z* = *z*.

**A2 Conditional exchangeability**: *A ⊥ T* (*z*) | *X* for *z* ∈ {0, 1}.

**A3 Positivity**: 0 *< P* (*Z* = *z* | *X*) *<* 1 almost surely.

**A4 Independent censoring**: *T ⊥ C* | *Z, X*.

**A5 Positivity of censoring**: *G*(*t* | *Z, X*) > 0 for *t ≤ τ*.

Under Assumptions 1-5, the causal restricted mean survival time (RMST) difference,

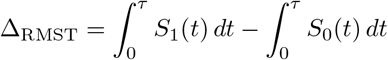

is identified from the observed data as

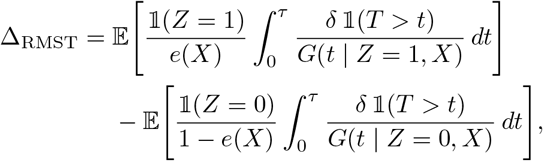

where *e*(*X*) = *P* (*Z* = 1 | *X*) is the propensity score, *G*(*t* | *Z, X*) = *P* (*C* > *t* | *Z, X*) is the conditional survival function for the censoring time, and 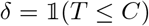 is the event indicator.

*Proof*. We will show that

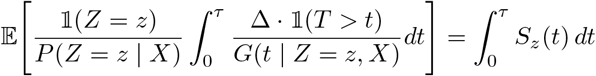

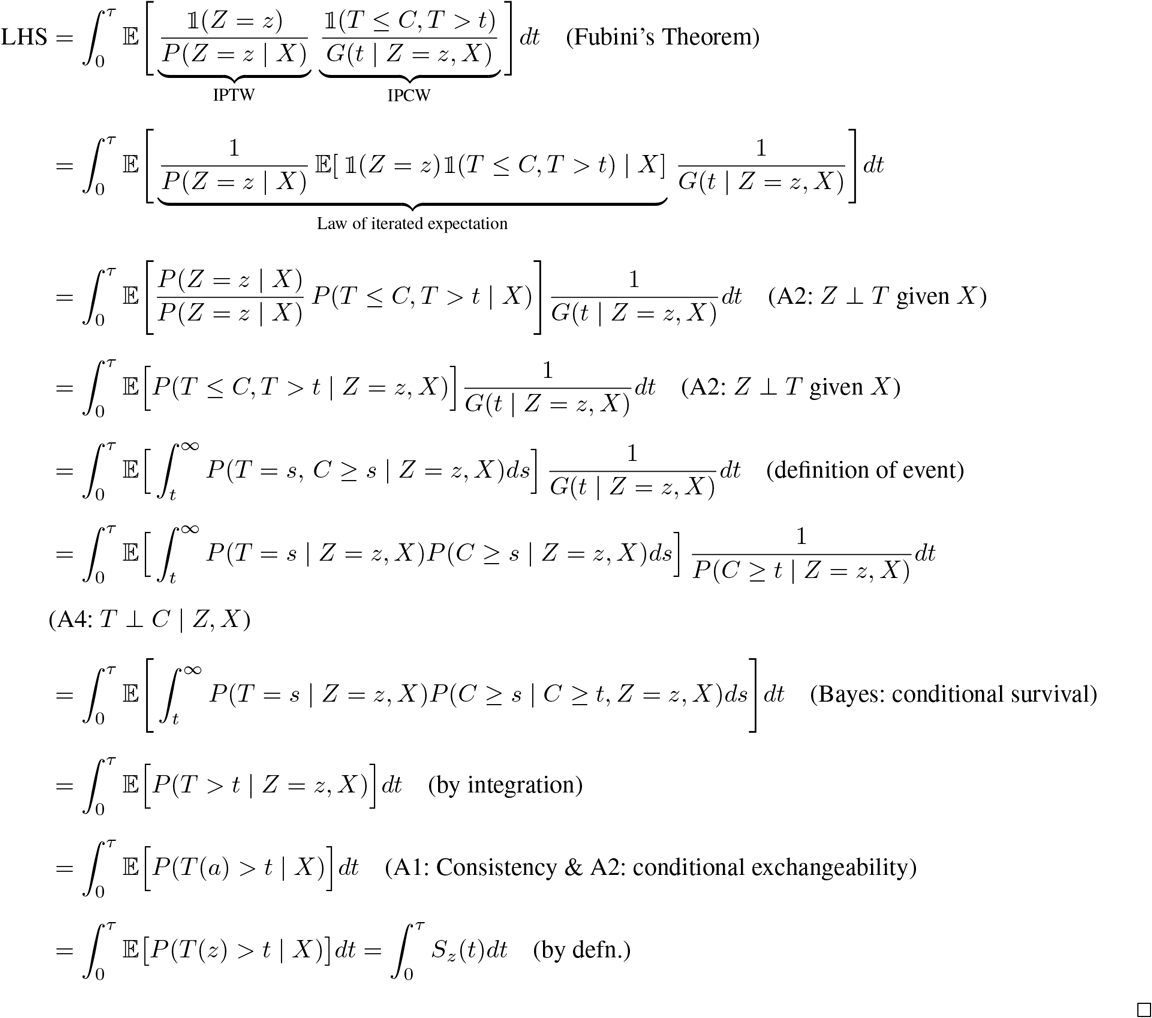

## Notes

### Competing Interest Statement

GH has received grant funding from Johnson & Johnson to support methods research not directly related to this study. Johnson & Johnson has not had input in the design, execution, interpretation of results or decision to publish. All other authors have no conflicting interests.

### Funding Statement

GH is funded by US National Institutes of Health grants (R01LM006910). The funders had no role in the design and conduct of the protocol; preparation, review, or approval of the manuscript; and decision to submit the manuscript for publication.

### Author Declarations

This study was approved by the Columbia University Institutional Review Board (approval date: December 2024; reference number AAAO7805).

